# Assessing the effect of medically assisted reproduction on hormone-related cancers in women: an emulated target trial

**DOI:** 10.1101/2025.11.16.25340364

**Authors:** Adrian Raymond Walker, Christos Venetis, Signe Opdal, Antoinette C Anazodo, Neville Hacker, Michael Chapman, Louisa Jorm, Robert J Norman, Catharyn Stern, Ursula M Sansom-Daly, Georgina Mary Chambers, Claire Melissa Vajdic

**Author notes:** Address correspondence to: Adrian Walker, Centre for Big Data in Health Research, Faculty of Medicine and Health, UNSW Sydney, NSW 2052, Australia. Joint first author (Adrian Raymond Walker, Christos Venetis). Joint senior author (Georgina Mary Chambers, Claire Melissa Vajdic).

## Abstract

1.

**Objective:** To determine the risk of hormone-related cancers following medically assisted reproductive treatment.

**Design:** Emulated target trial.

**Setting:** Australian health registries and administrative datasets.

**Participants:** 1,748,927 women enrolled in Medicare, Australia’s universal health insurance scheme, aged 18 to 55 between 1 January 1991 and 31 December 2018.

**Interventions:** Three exposures were defined from Medicare records of reimbursement for consultations, procedures and medications: assisted reproduction therapy (ART); intrauterine insemination/ovarian stimulation (IUI/OS); and ovulation induction with Clomiphene Citrate.

**Main outcome measures:** Hormone-related invasive cancers included breast, ovarian, uterine, thyroid, colorectal, and melanoma; in-situ cancers included breast and melanoma. Three cancers with no established hormonal links and high incidence – pancreatic, lung, and haematological – were included as negative controls. Flexible parametric survival models ascertained hazard ratios and cumulative marginal differences in incident cancers per 100,000 women. E-values assessed the risk of bias due to unmeasured confounding variables.

**Results:** Though most hormone-related cancers were elevated after MAR treatment (HRs: 1.09-1.64), E-value analysis suggested confounding due to underlying infertility conditions (endometriosis, polycystic ovarian syndrome) could account for this observed elevation for uterine, ovarian, and thyroid cancers. For any specific invasive cancer, fewer than 20 extra cancers per 100,000 women each year were predicted for treated versus comparator women. Emulated trials on the six hormone-related cancers showed increased cancer risk in the first years after treatment, however this was also observed for pancreatic and haematological cancers, suggesting detection bias. Haematological cancers showed an increased risk after treatment (HRs: 1.19-1.27), indicating uncontrolled confounding by ethnicity may account for the excess risk observed for haematological cancers and melanoma. Lung cancer risk was decreased after some treatments (HRs: 0.73-0.83).

**Conclusions:** Though MAR showed an association with some hormone-related cancers, the absolute difference in the number of expected cancers was small and may be explained by unmeasured confounding and detection bias.

**SUMMARY BOXES:** *Section 1: What is already known on this topic:*

- There is a biologically plausible link between medically assisted reproduction therapies and hormone-related cancers.
- Current evidence suggests exposure to medically assisted reproduction does not increase the risk of most hormone-related cancers.
- Assessments of causation in this context are challenging, as women undertaking medically assisted reproduction have a different hormone-related cancer risk profile to those who do not.

*Section 2: What this study adds:*

- This study is one of the largest examinations of women’s cancer risk following medically assisted reproduction and the first to use both the emulated target trial framework and negative control cancers, and to quantify the potential for unmeasured confounding.
- Most hormone-related cancers show a small elevation in relative and absolute risk following medically assisted reproduction therapies, though this may be due in part or whole to detection bias and unmeasured confounding from infertility-related conditions like endometriosis or polycystic ovary syndrome, anovulation, obesity, and ethnicity.
- This study provides a comprehensive assessment of the available evidence and guidance for improving future research on medically assisted reproduction and cancer.

## 3. INTRODUCTION

Medically assisted reproduction (MAR) refers to a collection of treatments that assist individuals to achieve a successful pregnancy. MAR treatments involving the extraction and in-vitro handling of ova (eggs) are referred to as *assisted reproductive technologies* (ART). These treatments are generally conducted with medical stimulation of the ovaries to retrieve multiple ova in a single menstrual cycle (known as *ovarian stimulation* or “OS”). Once the ova are retrieved, they are fertilised by either mixing them together with sperm (known as *in-vitro fertilisation* or “IVF”) or by selecting a single sperm and injecting it directly into an ovum (known as *intracytoplasmic sperm injection* or “ICSI”). If fertilisation is successful, the embryo is cultured in a laboratory and subsequently transferred into the uterus. MAR treatments without in-vitro handling of ova include *ovulation induction* (OI), where hormonal medications are administered to induce ovulation and achieve conception either by intercourse or by *intrauterine insemination* (IUI), where prepared sperm is inserted into the uterus around the time of ovulation. MAR treatments have become increasingly common, with around three million ART cycles performed each year worldwide.^1^

In most cases, MAR therapies involve the administration of medications that are either hormonal or hormone-modulating agents, including gonadotrophin-releasing hormone analogues, follicle stimulating hormone (FSH), human chorionic gonadotrophin, progesterone, oestrogen, and clomiphene citrate or letrozole. There has long been concern that these medicines might promote the development of hormone-related cancers, including ovarian, uterine, breast, melanoma, thyroid, and possibly colorectal cancers.^2–5^ In ART treatment, it is also possible the repeated puncture of follicles during ovum retrieval might cause breakdown and repair of the ovarian surface epithelium similar to that in natural ovulations, consistent with the ‘incessant ovulation hypothesis’ for ovarian carcinogenesis.^6, 7^ Despite the biological plausibility of these hypotheses,^6, 8–11^ strong evidence for any carcinogenic effect of MAR medicines is limited.^12, 13^

Understanding the risks of undertaking MAR treatments is critical for women and their healthcare practitioners.^14–20^ Due to the rarity of some cancers, the long latency period for most solid cancers, and ethical concerns around withholding treatment for infertility, randomised controlled trials cannot be used to examine this question.^12^ Therefore, the potential link between MAR and hormone-related cancers has only been explored using observational study designs, with inherent limitations in establishing causality. Such studies and meta-reviews have suggested there is little to no evidence that exposure to MAR treatments increases the risk of cancer.

This study aims to advance our knowledge of the relationship between MAR and hormone-related cancers. We leverage a population-based matched cohort of over 1.7 million women from Australia between 1991 and 2019, which represents one of the largest and longest population-based studies on cancer and MAR. Furthermore, we apply the emulated target trial approach, which aims to use observational data to mimic a randomised controlled trial by matching exposed and unexposed women as closely as possible at the time of study entry.^21^ To assess the potential for bias, we conduct an analysis of unmeasured confounding using E-values,^12, 22^ and include three non-hormone-related negative control cancers in our analysis. Finally, we use flexible parametric survival models^23^ to ascertain both the hazard ratio and absolute difference in predicted incident cancers over time, and to model a time-varying hazard ratio to assess for the potential of detection bias (i.e., cancer being more likely to be diagnosed while women are undergoing MAR procedures or while pregnant).^24, 25^

## 4. METHODS

### Data sources

The Australian Institute of Health and Welfare (AIHW) in conjunction with state-based Data Linkage Units conducted a bespoke linkage of jurisdictional and national population-based registries and administrative datasets (See Supplementary Table 1) to ascertain all variables. All data linkage was probabilistic. The AIHW used the Medicare Enrolment File as the linkage spine, and linked all other datasets to this spine based on individuals’ given name(s) and surname, phonetic encoding keys (double metaphone) for given name(s) and surname, sex, date of birth, residential postcode, geocoded residential address, and the first 6 letters of residential address.

### Target trial specification and emulation

We used the target trial emulation method to estimate the effect of exposure on cancer development, altering the exposure type and the cancer of interest ^21^. In brief, the method involves defining the ideal target trial for an intervention and then emulating that trial as closely as possible using observational data. Under strong assumptions, target trial emulation can estimate the true causal effect of a treatment on an outcome.^21, 26^ As well as the STROBE guidelines, we followed a draft version of the TARGET guidelines for reporting.^27, 28^ The optimal target trial is outlined in Supplementary Table 2 and shown below.

### Cohort eligibility

All people recorded as female in the Australian Medicare Enrolment File (MEF) who were alive and aged between 18 and 55 years at any point between 1 January 1991 and 31 December 2018 and who had no record in the MEF of ever living in Australia’s Northern Territory (due to lack of data availability) were eligible for inclusion. The MEF is a record of Australians registered to receive Medicare. Medicare is Australia’s public health care insurance scheme, of which all citizens and permanent residents are eligible, with almost all Australians registered soon after birth or residency. Data on these women were linked to the Medicare Benefits Schedule dataset (MBS; subsidised health services) and Pharmaceutical Benefits Scheme dataset (PBS; subsidised medicines) to identify medical consultations, procedures and medications for fertility treatments.

Initial selection of eligible treatment and comparator women was conducted by the AIHW. The AIHW partitioned women into “Exposed” (those who had received a MAR treatment) and “Unexposed” groups (those who had not).

To form the group of MAR-exposed eligible women, the AIHW identified all women in the inclusion period who were recorded in the MBS or PBS datasets as having received a relevant benefit for any MAR treatment (See Supplementary Table 3). Though MAR services and medicines can be accessed in Australia outside of Medicare, approximately 92% of ART treatments are reimbursed through the MBS.^29, 30^

To form the group of unexposed eligible women, each MAR-exposed woman was matched with up to four women with no record of receiving MAR treatment (matched randomly with replacement across exposed MAR women). Matching occurred on birth year, residential remoteness (metropolitan or rural), parity at matched exposed woman’s first MAR exposure date, and (if the matched exposed woman was parous at their first MAR) their age at their first birth +/- 365 days. All women in the unexposed group were alive on the date of the matched exposed woman’s first MAR exposure date.

To match exposed and unexposed women, it was necessary to estimate their parity over time using jurisdictional Perinatal Data Collections (PDCs). As the starting date for each jurisdictional PDC data varied (Supplementary Table 1), women were only eligible for inclusion and matching once PDC data became available in their State.

Following cohort provision by the AIHW, further data cleaning steps were undertaken by researchers. Figure 1 shows the steps taken to ascertain the cohort of eligible women for treatment strategy assignment.

**Figure 1:**
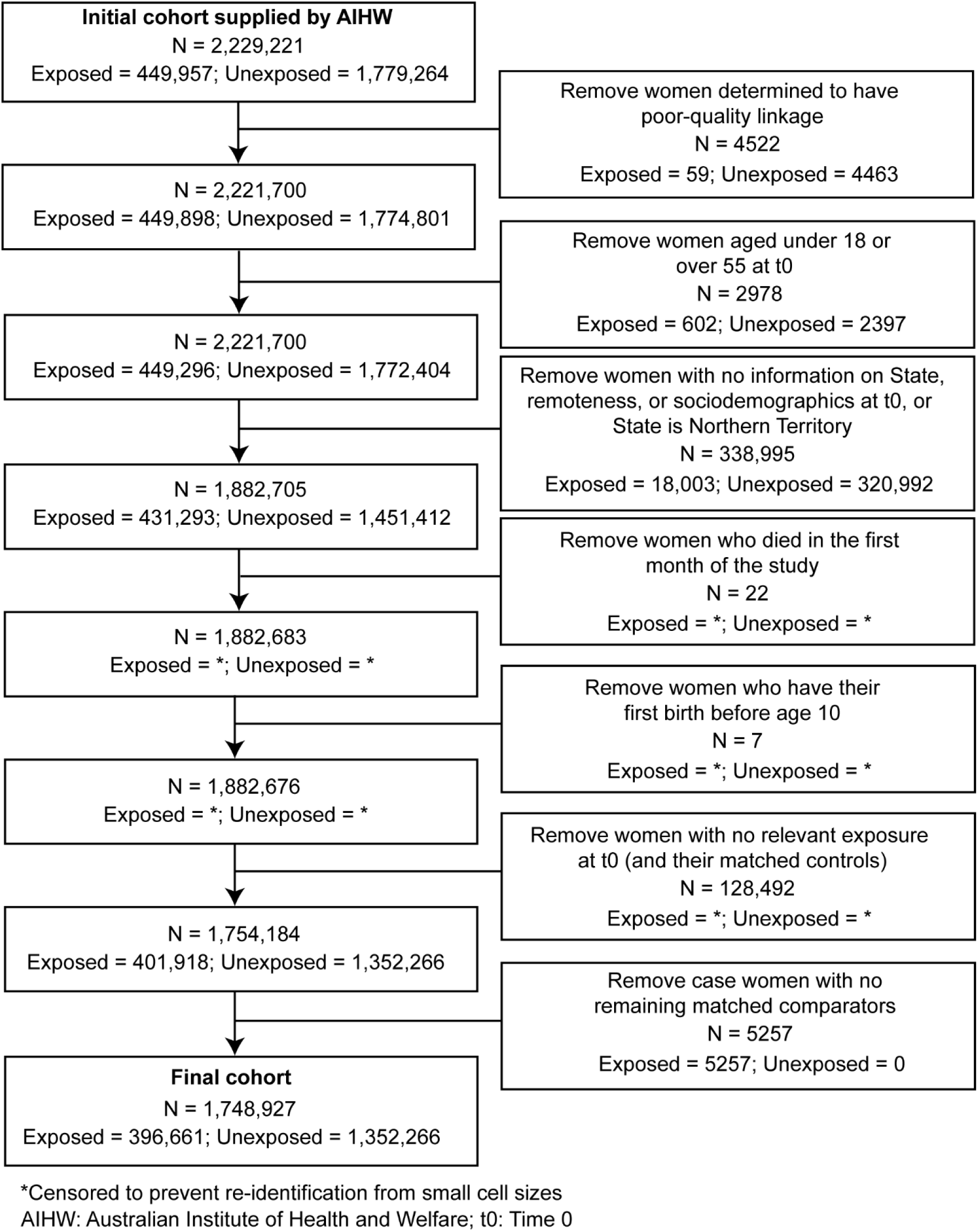
Construction of eligible exposed and unexposed women, prior to partitioning to treatment comparator cohorts

### Treatment strategies

Three different types of MAR treatment were identified by the presence of relevant MBS and/or PBS codes (Supplementary Table 4):

1. ART treatments (IVF or ICSI) (hereafter “ART”);
2. Intrauterine insemination (IUI) with ovarian stimulation with FSH *or* ART cancelled before egg retrieval (hereafter “IUI/OS”); and
3. Ovulation induction using Clomiphene Citrate (hereafter “Clomiphene Citrate”).

### Assignment procedure

Women were classified into treatment groups was conducted based the criteria presented in Supplementary Table 4. Assignment was based on specific MBS or PBS codes within 28 days of first MAR exposure. Overall, 93.4% of women identified as MAR-exposed by AIHW were classified into a treatment group. The remaining women did not have treatment codes relevant for the study in the allotted time and were excluded from the study. For example, this may have occurred if a woman was dispensed a MAR drug (other than Clomiphene Citrate), but there was no MBS reimbursement recorded within 28 days for initiating a MAR treatment (e.g., due to falling pregnant after filling the prescription but before starting treatment, or receiving treatment without MBS reimbursement).

A woman was assigned to the respective comparator group if she was matched to the MAR-exposed woman selected for inclusion by the AIHW. As prescription data only became available from 1 July 2002, women were only assigned to the emulated trial on Clomiphene Citrate from this date.

### Follow-up

Data were partitioned into 28-day windows for analysis, starting from the day of first recorded MBS or PBS code indicating MAR treatment or matched day for the comparator group. This approach was taken to approximate the length of an IVF treatment cycle, with each year approximated as 364 days (13 cycles).

Time zero (t0) was defined as the first 28-day window in which treatment assignment occurred, with follow-up beginning from the next 28-day window.

Women were followed up until either a relevant cancer outcome, 31 December 2019, or death (as recorded in the National Death Index), whichever occurred first.

### Outcome definition

Hormone-related cancers were chosen as outcomes, comprising breast (invasive, in-situ), ovarian (serous, non-serous, any), uterine, melanoma (invasive, in-situ), colorectal, and thyroid cancer, identified by a record in the Australian Cancer Database (ACD). Cancers not considered hormone-related (pancreatic, lung, and haematological) were included as negative controls. Only invasive tumours were considered (International classification of disease – Oncology 3^rd^ edition: behaviour code 3), unless specified. All cancers were classified using ICD-O3 topographies and morphologies (Supplementary Table 5).

### Causal contrasts

All emulated trials used an intention-to-treat protocol.

### Identifying assumptions

It was assumed that available variables appropriately controlled for residual confounding, and the likelihood of censoring did not systematically differ between those assigned to the treatment and comparator groups.

### Statistical analysis

For each target trial, stabilized inverse propensity score weighting was used to control for confounding.^31^ Weights were estimated using logistic regression, predicting the likelihood of assignment to the treatment group from the confounding set.

Confounding variables included:

- Age at t0;
- History of giving birth prior to t0;
- Residential remoteness at t0 (metropolitan, inner regional, remote/very remote);
- Average area-based Index of Relative Socioeconomic Disadvantage percentile, based on available lifetime data for Australian area of residence;
- Subsidised diabetes medication record prior to t0; and
- Cancer registry notified cancer diagnosis prior to t0.

Confounding variables included in our study were determined based on the directed acyclic graph in Walker et a1.^12^ and the available data.

Following weighting, four nested flexible parametric survival models^23^ with robust variance estimators were run. To account for similarities between treated women and their matched comparators, we used cluster-robust variance estimators, treating each MAR-treated woman and her matched comparators as a cluster. To determine whether a time-varying effect was required to model the hazard function of treatment and, if so, the number of required splines, four models were compared using likelihood ratio tests, preferring simpler models due to parsimony (non-time-varying, 0 spline, 1 spline, and 3 spline). Further description of the survival modelling and selection process is given in Supplementary Methods 1.

For emulated trials where the non-time-varying model was preferred, the mean hazard ratio across the full follow-up period is reported. For emulated trials where a time-varying effect of treatment was preferred, the hazard ratio changes over time are shown graphically.

For each model, the cumulative marginal difference in incident cancers between the treatment and comparator groups over time was estimated and reported as the number of additional cancer cases expected per 100,000 women at year 1, and then every five years starting at year 5. This was done using the *standsurv* postestimation command in STATA.^32^ Briefly, the command models the average survival curves for the entire population assuming either treatment or non-treatment, controlling for the included confounding variables. By comparing the marginal survival curve assuming treatment to the marginal survival curve assuming non-treatment at different times from t0, one can determine how many extra incident cancers were predicted after a certain number of years. This process is similar to comparing Kaplan-Meier survival curves, but crucially provides an estimate of the survival difference adjusted for available confounding variables.

In each emulated trial, E-values^22^ were calculated to assess the likelihood uncontrolled confounding impacted our findings. Briefly, E-values are measured on the risk-ratio scale and reflect the weakest association between an uncontrolled confounder and both the exposure and outcome, that could fully explain the observed effect. A full discussion of E-values and their use in studies of MAR treatments and health outcomes is given in Walker, Venetis (12) When hazard ratios changed over time, the E-value was assessed at year 1, and then every five years starting at year 5.

We conducted a complete case analysis of the data. We assumed data were missing at random, that is, that missingness in the outcome (due to either lack of detection during the study period or migration out of the country) were dependent on our observed variables (particularly age, remoteness, and socioeconomic status). Analysis was conducted using SAS version 9 and STATA 18.

### Study registration

This study, including rationale, target trial specification, and analysis plan, was pre-registered on the Open Science Framework at https://osf.io/rk9n6.^33^ Although the study was pre-registered, some elements of the design and analysis were adapted during implementation to strengthen feasibility and validity. These modifications and their rationale are described in detail in Supplementary Methods 2.

### Ethics approval

The study was approved by all relevant human research ethics committees (HRECs) including the AIHW HREC (EO2019/5/1061). Data was accessed and used under a waiver of informed consent. Researchers were granted access to linked anonymised data.

### Patient and public involvement statement

A consumer with lived-experience was a member of the project advisory group, bringing perspectives on the priorities and experiences of women who use MAR. The consumer reviewed this manuscript before submission and provided input to ensure the messaging was clear and appropriate for a broad audience. A broader group of MAR consumers will assist with the development of project briefs for the public and healthcare sector.

## 5. RESULTS

### Demographics

Table 1 shows the demographic profile of the three MAR treatment groups and their comparators, and Supplementary Table 6 shows the total number of incident cancers. Generally, the comparator cohorts were well-matched to the relevant treatment cohort. Those receiving MAR treatments were more likely to live in less disadvantaged areas and areas with high levels of education and employment. The number of women receiving MAR treatment increased over time. Most individuals were nulliparous when entering the cohort. Due to an error in data extraction, a substantial proportion of comparator individuals (around 300,000) identified by the AIHW for study inclusion were not in the country at t0 and had to be excluded, as we could not determine the presence or absence of cancer. This error created an imbalance in the number of nulliparous women in the MAR treatment and comparator groups, which was accounted for later through inverse probability of treatment weighting.

**Table 1:** Demographic profile of the three treatment and three comparator cohorts. Percentages are taken column-wise (except for total *N*, given as percentage across treatment type)

|  | ART treatment |  | IUI/OS treatment |  | Clomid dispensation |  |
| --- | --- | --- | --- | --- | --- | --- |
|  | Treatment | Comparator | Treatment | Comparator | Treatment | Comparator |
| **N** | 171,609 (22.7%) | 584,311 (77.3%) | 77,767 (23.0%) | 260,280 (77.0%) | 156,084 (22.5%) | 537,796 (77.5%) |
| **Age at study entry** |  |  |  |  |  |  |
| Mean (SD) | 34.669 (5.054) | 34.851 (5.022) | 34.019 (5.143) | 34.233 (5.116) | 31.766 (5.434) | 32.008 (5.442) |
| Median [IQR] | 35 [31, 38] | 35 [31, 39] | 34 [30, 38] | 34 [31, 38] | 31 [28, 35] | 32 [28, 35] |
| **Year at t0** |  |  |  |  |  |  |
| 1991-1994 | 12,540 (7.3%) | 37,985 (6.5%) | 8,486 (10.9%) | 25,621 (9.8%) | N/A | N/A |
| 1995-1999 | 17,139 (10.0%) | 54,633 (9.3%) | 13,438 (17.3%) | 42,572 (16.4%) | N/A | N/A |
| 2000-2004 | 22,464 (13.1%) | 75,113 (12.9%) | 13,185 (17.0%) | 43,654 (16.8%) | 25,671 (16.4%) | 86,458 (16.1%) |
| 2005-2009 | 36,233 (21.1%) | 126,562 (21.7%) | 13,854 (17.8%) | 48,365 (18.6%) | 47,008 (30.1%) | 162,948 (30.3%) |
| 2010-2014 | 43,925 (25.6%) | 151,406 (25.9%) | 16,002 (20.6%) | 55,099 (21.2%) | 51,456 (33.0%) | 175,471 (32.6%) |
| 2015-2018 | 39,308 (22.9%) | 138,612 (23.7%) | 12,802 (16.5%) | 44,969 (17.3%) | 31,949 (20.5%) | 112,919 (21.0%) |
| **State** |  |  |  |  |  |  |
| ACT | 2,861 (1.7%) | 10,042 (1.7%) | 2,144 (2.8%) | 4,659 (1.8%) | 2,591 (1.7%) | 9,272 (1.7%) |
| NSW | 62,546 (36.4%) | 197,861 (33.9%) | 24,527 (31.5%) | 90,755 (34.9%) | 53,463 (34.3%) | 178,278 (33.1%) |
| QLD | 26,869 (15.7%) | 110,667 (18.9%) | 20,631 (26.5%) | 47,510 (18.3%) | 33,378 (21.4%) | 104,871 (19.5%) |
| SA | 12,779 (7.4%) | 35,766 (6.1%) | 4,287 (5.5%) | 15,806 (6.1%) | 9,263 (5.9%) | 35,336 (6.6%) |
| TAS | 2,985 (1.7%) | 11,154 (1.9%) | 1,673 (2.2%) | 4,994 (1.9%) | 2,707 (1.7%) | 10,996 (2.0%) |
| VIC | 48,565 (28.3%) | 161,767 (27.7%) | 15,910 (20.5%) | 71,824 (27.6%) | 45,598 (29.2%) | 143,750 (26.7%) |
| WA | 15,004 (8.7%) | 57,054 (9.8%) | 8,595 (11.1%) | 24,732 (9.5%) | 9,084 (5.8%) | 55,293 (10.3%) |
| **Remoteness** |  |  |  |  |  |  |
| Major cities | 141,187 (82.3%) | 463,758 (79.4%) | 64,901 (83.5%) | 207,410 (79.7%) | 121,390 (77.8%) | 418,872 (77.9%) |
| Inner regional | 20,091 (11.7%) | 82,804 (14.2%) | 9,201 (11.8%) | 37,701 (14.5%) | 23,064 (14.8%) | 78,463 (14.6%) |
| Outer regional | 8,544 (5.0%) | 31,639 (5.4%) | 3,213 (4.1%) | 12,683 (4.9%) | 9,740 (6.2%) | 33,592 (6.2%) |
| Remote | 1,234 (0.7%) | 3,946 (0.7%) | 317 (0.4%) | 1,636 (0.6%) | 1,379 (0.9%) | 4,492 (0.8%) |
| Very remote | 553 (0.3%) | 2,164 (0.4%) | 135 (0.2%) | 850 (0.3%) | 511 (0.3%) | 2,377 (0.4%) |
| **Index of Relative Socioeconomic Disadvantage quintile** |  |  |  |  |  |  |
| 1 | 20,996 (12.2%) | 86,587 (14.8%) | 9,191 (11.8%) | 38,582 (14.8%) | 26,832 (17.2%) | 83,173 (15.5%) |
| 2 | 22,700 (13.2%) | 88,645 (15.2%) | 9,724 (12.5%) | 39,248 (15.1%) | 24,525 (15.7%) | 85,919 (16.0%) |
| 3 | 31,465 (18.3%) | 110,299 (18.9%) | 14,638 (18.8%) | 48,816 (18.8%) | 31,876 (20.4%) | 105,123 (19.5%) |
| 4 | 37,234 (21.7%) | 123,099 (21.1%) | 18,029 (23.2%) | 54,531 (21.0%) | 32,462 (20.8%) | 112,598 (20.9%) |
| 5 | 59,214 (34.5%) | 175,681 (30.1%) | 26,185 (33.7%) | 79,103 (30.4%) | 40,389 (25.9%) | 150,983 (28.1%) |
| **Estimated parity at t0** |  |  |  |  |  |  |
| 0 | 138,542 (80.7%) | 420,180 (71.9%) | 62,704 (80.6%) | 190,969 (73.4%) | 107,942 (69.2%) | 339,679 (63.2%) |
| 1 | 20,649 (12.0%) | 70,216 (12.0%) | 10,853 (14.0%) | 32,053 (12.3%) | 34,391 (22.0%) | 97,530 (18.1%) |
| 2 | 7,363 (4.3%) | 54,922 (9.4%) | 2,813 (3.6%) | 21,946 (8.4%) | 9,100 (5.8%) | 62,385 (11.6%) |
| 3 | 3,182 (1.9%) | 23,990 (4.1%) | 898 (1.2%) | 9,500 (3.6%) | 2,880 (1.8%) | 24,809 (4.6%) |
| 4 | 1,253 (0.7%) | 9,046 (1.5%) | 308 (0.4%) | 3,478 (1.3%) | 1,020 (0.7%) | 8,685 (1.6%) |
| 5 and above | 620 (0.4%) | 5,957 (1.0%) | 191 (0.2%) | 2,334 (0.9%) | 751 (0.5%) | 4,708 (0.9%) |
| **History of diabetes at t0** |  |  |  |  |  |  |
| Yes | 3,876 (2.3%) | 8,431 (1.4%) | 1,583 (2.0%) | 2,893 (1.1%) | 10,776 (6.9%) | 8,109 (1.5%) |
| **ART\*** |  |  |  |  |  |  |
| At t0 | 171,609 (100.0%) | N/A | 1,715 (2.2%) | N/A | 309 (0.2%) | N/A |
| Total number of treatments | 510,169 | N/A | 32,052 | N/A | 18,221 | N/A |
| **IUI/OS\*** |  |  |  |  |  |  |
| At t0 | 1,715 (1.0%) | N/A | 77,767 (100.0%) | N/A | 6,797 (4.4%) | N/A |
| Total number of treatments | 161,166 | N/A | 213,646 | N/A | 29,760 | N/A |
| **Clomiphene Citrate\*** |  |  |  |  |  |  |
| At t0 | 309 (0.2%) | N/A | 6,797 (8.7%) | N/A | 156,084 (100.0%) | N/A |
| Total number of treatments | 150,197 | N/A | 68,023 | N/A | 439,388 | N/A |
| **Years in study** |  |  |  |  |  |  |
| Mean (SD) | 11.083 (7.473) | 10.781 (7.319) | 13.439 (7.894) | 13.046 (7.807) | 9.083 (4.654) | 9.029 (4.665) |
| Median [IQR] | 10 [5, 16] | 10 [5, 16] | 13 [6, 20] | 12 [6, 20] | 9 [5, 13] | 9 [5, 13] |
| **Died in study period** |  |  |  |  |  |  |
| Yes | 1,512 (0.9%) | 5,824 (1.0%) | 777 (1.0%) | 3,318 (1.3%) | 627 (0.4%) | 2,610 (0.5%) |

\*When approximating the number of treatments, only one treatment could occur in each 28-day analysis window. IQR=interquartile range; SD=standard deviation; t0=time zero; ART=Assisted Reproductive Technology; IUI/OS=Intrauterine Insemination/Ovarian Stimulation

Mean follow-up time was roughly equal in all emulated trials (ART 11.1 years, comparator 10.8 years; IUI/OS 13.4 years, comparator 13.0 years; Clomiphene Citrate 9.1 years, comparator 9.0 years).

### Survival modelling

#### Model selection

All results of the model selection step can be seen in Supplementary Table 7. Generally, models with constant hazard ratios were preferred. Models with time-varying hazard ratios were preferred in eight comparisons of hormone-related cancers: for ART this included ovarian (all and serous; 0 splines), uterine (0 splines), invasive melanoma (3 splines); for IUI/OS, this included invasive melanoma (0 splines), and non-serous ovarian (3 splines); and for Clomiphene Citrate, this included invasive breast (1 spline) and uterine cancers (0 splines). Models with time-varying hazard were also preferred for two negative control cancers (pancreatic, 0 splines; haematological, 0 spline) following ART.

#### Hazard ratios

Table 2 and Supplementary Table 8 show the hazard ratios, associated E-values, and cumulative marginal totals and differences in cancer incidence per 100,000 women for the treatment compared to the comparator group for each emulated trial. Figure 2 shows that these findings for cancers that varied over time.

**Figure 2:**
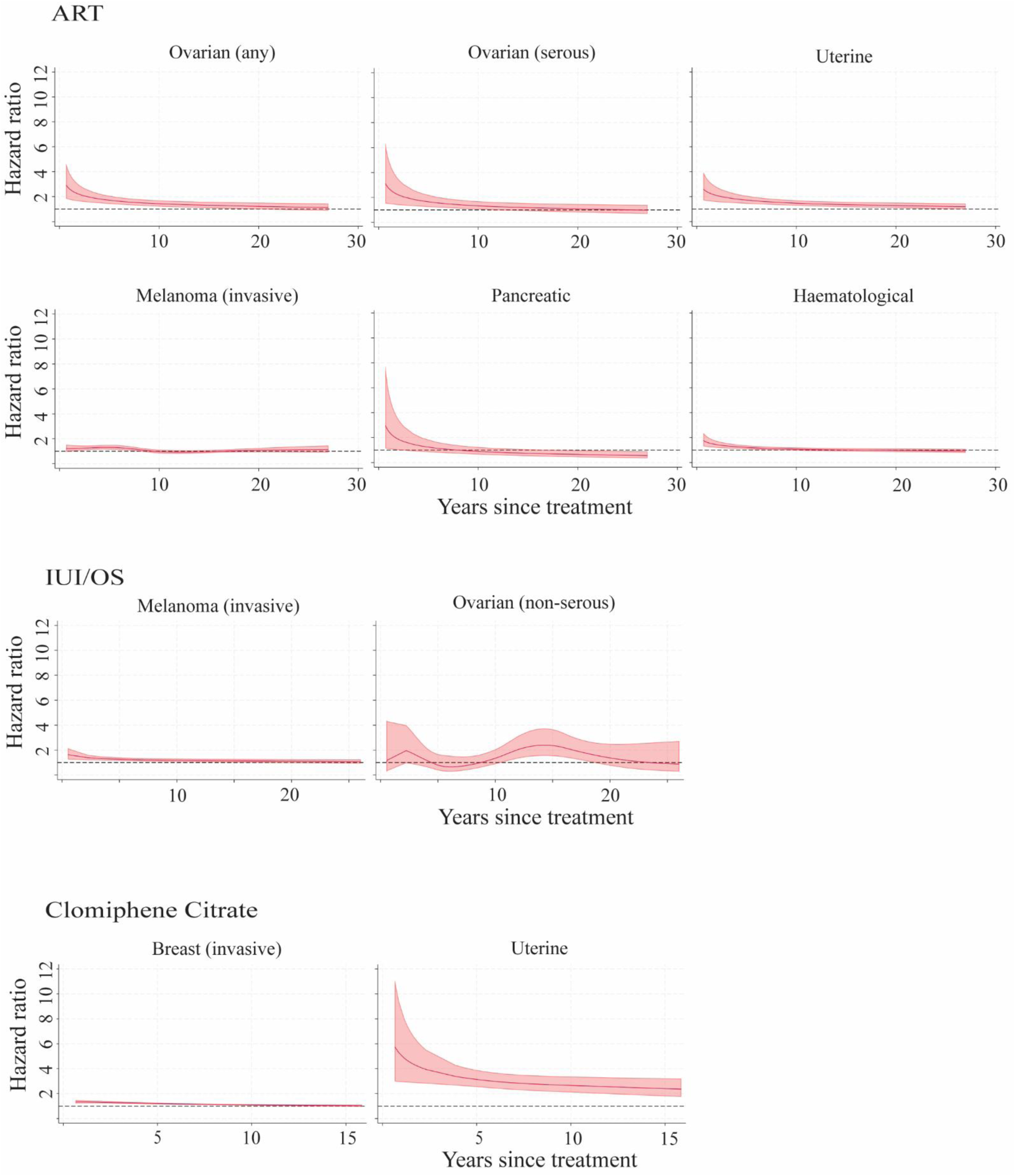
Hazard ratios over time for cancers with time-varying associations with MAR treatments. es start from 6 months to improve figure readability.

**Table 2:** Hazard ratios, E-values, and cumulative marginal difference in incident cancers (per 100,000 women) for each emulated target trial.

| MAR treatment type | Hazard ratio |  |  | Cumulative marginal difference in incident cancers per 100,000 women |  |  |  |  |  |
| --- | --- | --- | --- | --- | --- | --- | --- | --- | --- |
| Cancer type | Estimate<br>(95% CI) | Stand-<br>ard<br>error | E-Value<br>(95% CI) | 1 year | 5 years | 10 years | 15 years | 20 years | 25 years |
| ART |  |  |  |  |  |  |  |  |  |
| Breast (invasive) | 1.11 (1.06, 1.16) | 0.03 | 1.45 (1.30, 1.59) | 3 (2, 5) | 31 (16, 45) | 94 (51, 138) | 187 (101, 274) | 303 (163, 443) | 438 (236, 640) |
| Breast (in-situ) | 1.30 (1.10, 1.52) | 0.11 | 1.91 (1.44, 2.42) | 1 (0, 2) | 12 (4, 19) | 43 (14, 71) | 98 (32, 164) | 180 (58, 301) | 286 (90, 482) |
| Ovarian | Time varying |  |  |  |  | 5 (2, 7) | 19 (11, 27) | 36 (22, 51) | 54 (31, 77) |
| Ovarian (serous) | Time varying |  |  |  |  | 2 (0, 3) | 8 (3, 13) | 15 (5, 24) | 21 (5, 37) |
| Ovarian (non-serous) | 1.64 (1.31, 2.05) | 0.19 | 2.66 (1.94, 3.52) | 1 (0, 2) | 7 (3, 11) | 17 (8, 27) | 32 (15, 49) | 53 (26, 80) | 79 (38, 120) |
| Uterine | Time varying |  |  |  |  | 4 (2, 6) | 16 (9, 24) | 37 (23, 51) | 64 (40, 88) |
| Thyroid | 1.16 (1.04, 1.29) | 0.06 | 1.58 (1.23, 1.90) | 2 (0, 3) | 13 (3, 22) | 28 (6, 50) | 47 (10, 84) | 69 (15, 123) | 94 (21, 167) |
| Colorectal | 1.12 (1.00, 1.25) | 0.06 | 1.48 (1.02, 1.80) | 1 (0, 2) | 6 (0, 12) | 16 (0, 33) | 32 (-1, 65) | 56 (-1, 114) | 90 (-2, 182) |
| Melanoma (invasive) | Time varying |  |  |  |  | 6 (-2, 13) | 39 (16, 62) | 67 (29, 105) | 51 (-4, 107) |
| Melanoma (in-situ) | 1.14 (1.03, 1.27) | 0.06 | 1.54 (1.19, 1.85) | 4 (1, 7) | 24 (4, 44) | 66 (11, 120) | 138 (24, 252) | 235 (40, 430) | 353 (59, 648) |
| Pancreatic | Time varying |  |  |  |  | 1 (0, 3) | 3 (-1, 7) | 3 (-4, 10) | -1 (-13, 10) |
| Lung | 0.82 (0.69, 0.97) | 0.07 | * | 0 (-1, 0) | -3 (-5, 0) | -10 (-18, -2) | -23 (-42, -4) | -48 (-86, -9) | -91 (-165, -17) |
| Haematological cancer | Time varying |  |  |  |  | 6 (2, 10) | 22 (9, 34) | 33 (12, 53) | 38 (5, 71) |
| IUI/OS |  |  |  |  |  |  |  |  |  |
| Breast (invasive) | 1.09 (1.03, 1.16) | 0.03 | 1.40 (1.19, 1.59) | 3 (1, 5) | 24 (7, 42) | 75 (21, 128) | 151 (43, 258) | 246 (71, 422) | 358 (103, 613) |
| Breast (in-situ) | 1.16 (0.90, 1.51) | 0.15 | * | 1 (-1, 2) | 8 (-6, 21) | 24 (-19, 66) | 49 (-38, 136) | 83 (-64, 230) | 125 (-96, 346) |
| Ovarian | 1.48 (1.20, 1.84) | 0.16 | 2.33 (1.68, 3.08) | 1 (0, 3) | 9 (3, 15) | 25 (10, 41) | 48 (19, 78) | 81 (32, 130) | 125 (50, 200) |
| Ovarian (serous) | 1.49 (1.10, 2.04) | 0.24 | 2.35 (1.42, 3.49) | 0 (0, 1) | 3 (0, 6) | 10 (1, 18) | 21 (3, 39) | 40 (6, 74) | 69 (10, 127) |
| Ovarian (non-serous) | Time varying |  |  |  |  | 0 (-3, 3) | 6 (-5, 17) | 4 (-12, 19) | 29 (3, 56) |
| Uterine | 1.54 (1.29, 1.84) | 0.14 | 2.45 (1.90, 3.09) | 1 (0, 2) | 9 (4, 13) | 28 (15, 42) | 63 (33, 93) | 124 (66, 181) | 223 (120, 325) |
| Thyroid | 1.11 (0.95, 1.29) | 0.09 | * | 1 (-1, 3) | 8 (-4, 21) | 19 (-10, 48) | 31 (-16, 79) | 46 (-23, 114) | 61 (-32, 154) |
| Colorectal | 0.99 (0.86, 1.15) | 0.08 | * | 0 (-1, 1) | 0 (-8, 7) | -1 (-22, 20) | -2 (-41, 38) | -3 (-73, 67) | -4 (-118, 109) |
| Melanoma (invasive) | Time varying |  |  |  |  | 17 (6, 28) | 61 (29, 92) | 99 (52, 147) | 135 (66, 204) |
| Melanoma (in-situ) | 1.39 (1.19, 1.64) | 0.11 | 2.14 (1.66, 2.66) | 7 (3, 11) | 59 (28, 91) | 173 (81, 264) | 352 (165, 538) | 584 (269, 900) | 864 (389, 1340) |
| Pancreatic | 0.85 (0.57, 1.27) | 0.17 | * | 0 (0, 0) | -1 (-3, 1) | -2 (-8, 3) | -5 (-18, 7) | -11 (-36, 15) | -21 (-70, 29) |
| Lung | 0.72 (0.57, 0.91) | 0.09 | * | 0 (-1, 0) | -4 (-7, -1) | -14 (-24, -5) | -33 (-55, -11) | -68 (-112, -23) | -130 (-215, -45) |
| Haematological cancer | 1.27 (1.11, 1.46) | 0.09 | 1.86 (1.45, 2.28) | 3 (1, 4) | 16 (6, 26) | 38 (14, 62) | 69 (27, 112) | 119 (46, 192) | 190 (73, 308) |
| **Clomiphene Citrate** |  |  |  |  |  |  |  |  |  |
| Breast (invasive) |  | Time varying |  | 9 (2, 17) | 64 (37, 91) | 104 (51, 157) | 64 (-45, 174) | N/A | N/A |
| Breast (in-situ) | 1.15 (0.97, 1.38) | 0.10 | * | 0 (0, 1) | 3 (-1, 8) | 14 (-4, 31) | 34 (-9, 76) | N/A | N/A |
| Ovarian | 1.40 (1.07, 1.81) | 0.19 | 2.14 (1.36, 3.03) | 1 (0, 2) | 7 (1, 13) | 16 (2, 29) | 27 (4, 50) | N/A | N/A |
| Ovarian (serous) | 1.36 (0.90, 2.06) | 0.29 | * | 0 (0, 1) | 2 (-1, 5) | 6 (-3, 14) | 11 (-5, 26) | N/A | N/A |
| Ovarian (non-serous) | 1.42 (1.01, 1.99) | 0.24 | 2.19 (1.12, 3.38) | 1 (0, 2) | 5 (0, 10) | 10 (-1, 20) | 16 (-1, 34) | N/A | N/A |
| Uterine |  | Time varying |  | 6 (3, 9) | 33 (23, 43) | 75 (58, 93) | 144 (107, 180) | N/A | N/A |
| Thyroid | 1.30 (1.15, 1.46) | 0.08 | 1.91 (1.55, 2.29) | 3 (1, 5) | 24 (12, 36) | 58 (29, 88) | 96 (47, 144) | N/A | N/A |
| Colorectal | 1.15 (0.99, 1.35) | 0.09 | * | 1 (0, 2) | 7 (-1, 16) | 19 (-2, 40) | 37 (-4, 79) | N/A | N/A |
| Melanoma (invasive) | 1.22 (1.10, 1.34) | 0.06 | 1.73 (1.44, 2.02) | 4 (2, 7) | 29 (14, 44) | 67 (32, 103) | 120 (57, 183) | N/A | N/A |
| Melanoma (in-situ) | 1.29 (1.17, 1.42) | 0.06 | 1.91 (1.63, 2.20) | 5 (3, 7) | 37 (22, 51) | 106 (63, 149) | 237 (142, 333) | N/A | N/A |
| Pancreatic | 0.96 (0.57, 1.60) | 0.25 | * | 0 (0, 0) | 0 (-2, 1) | -1 (-6, 5) | -1 (-15, 13) | N/A | N/A |
| Lung | 0.95 (0.72, 1.26) | 0.14 | * | 0 (0, 0) | -1 (-4, 3) | -2 (-11, 8) | -5 (-31, 22) | N/A | N/A |
| Haematological cancer | 1.18 (1.02, 1.37) | 0.09 | 1.65 (1.18, 2.07) | 2 (0, 3) | 10 (1, 20) | 26 (2, 49) | 47 (5, 90) | N/A | N/A |

\*E-value not calculated for results where hazard ratio confidence interval is entirely below or includes 1
ART=Assisted Reproductive Technology; IUI/OS=Intrauterine Insemination/Ovarian Stimulation

For emulated trials on hormone-related cancers without a time-varying hazard ratio, cancer risk was generally elevated following all types of MAR treatment, with hazard ratio estimates ranging from 1.09 (invasive breast cancer following IUI/OS) to 1.64 (non-serous ovarian cancer following ART). In general, for emulated trials suggesting a time-constant hazard ratio, invasive breast cancer (hazard ratios ranging 1.09 to 1.11) had the weakest association, while any ovarian (hazard ratios ranging 1.40 to 1.48) and uterine cancer (1.54, 95% confidence interval 1.29 to 1.84) had the strongest. The exceptions to an elevation in risk were in-situ breast (1.16, 0.90 to 1.51), thyroid (1.11, 0.95 to 1.29), and colorectal cancer (0.99, 0.86 to 1.15) following IUI/OS, and in-situ breast (1.15, 0.97 to 1.38), serous ovarian cancer (1.36, 0.90 to 2.06), and colorectal cancer (1.15, 0.99-1.35) following Clomiphene Citrate use.

Eight emulated trials of hormone-related cancers showed evidence of a time-varying hazard ratio, and the most common pattern was an increased risk immediately post first treatment that decreased over time, reducing to a small or no difference in risk after around 10 years.

This was the case for any ovarian cancer, serous ovarian cancer (47% of ovarian cancers), uterine cancer following ART, invasive melanoma following IUI/OS, and invasive breast and uterine cancer following Clomiphene Citrate. The excess cancer risk was maintained across the follow-up period for only one cancer-exposure combination, uterine cancer following Clomiphene Citrate. Invasive melanoma risk following ART and non-serous ovarian cancer following IUI/OS showed a different risk pattern over time, with increasing risk up to around five years post treatment, and then a decrease, reaching no excess risk before 10 years post treatment.

Pancreatic cancer showed no association with IUI/OS and Clomiphene Citrate, and an increased risk following ART. This association with ART was time-varying, with increased risk in the five years after treatment that decreased over time, a pattern that mirrored most hormone-related cancers with a time-varying hazard ratio. A similar pattern was observed for haematological cancers following ART, however there was also an increased risk of these cancers following IUI/OS (1.27, 1.11 to 1.46) and Clomiphene Citrate (1.18, 1.02 to 1.37). Lung cancer risk was decreased after ART (0.82, 0.69 to 0.97) and IUI/OS (0.72, 0.57 to 0.91) and there was no association with Clomiphene Citrate (0.95, 0.72 to 1.26).

#### Cumulative marginal difference in incident cancers

Uterine cancer, in-situ melanoma, and invasive breast cancer showed a notable difference in the cumulative predicted numbers of incident cancers per 100,000 women for treated compared to untreated women (Table 2, Supplementary Table 8). The absolute risks of uterine cancer and in-situ melanoma were increased for all treatment types at 25 years after treatment (15 years for Clomiphene Citrate due to the shorter follow-up): 159 excess uterine cancers (95% confidence interval 72 to 246) and 353 excess in-situ melanomas (59 to 648) were predicted per 100,000 women after ART; 223 excess uterine cancers (120 to 325) and 864 excess in-situ melanomas (389 to 1340) were predicted per 100,000 women after IUI/OS; and 144 excess uterine cancers (107 to 180) and 237 (142 to 333) in-situ melanoma were predicted per 100,000 women after Clomiphene Citrate. The number of invasive breast cancers was elevated per 100,000 women following ART (438, 236 to 640) and IUI/OS (358, 103 to 613) after 25 years, but for Clomiphene Citrate, the cumulative marginal difference reduced after 10 years, such that by 15 years no extra cancers were predicted (64, −45 to 174).

Invasive melanoma also accounted for a substantial number of cancers per 100,000 women 25 years after IUI/OS (205, 56 to 354) and 15 years after Clomiphene Citrate (120, 57 to 183). For ART, growth in the cumulative marginal difference slowed substantially after 10 years, such that by 25 years no extra cancers were predicted (102, −32 to 236).

Though all treatments showed a strong signal for a relationship with ovarian cancer, due to the smaller number of ovarian cancers in the population, the overall cumulative marginal difference in ovarian cancer per 100,000 women between treatment and comparator groups was relatively small compared to other types of cancers (ART after 25 years: 94, 31 to 156; IUI/OS after 25 years: 125, 50 to 200; Clomiphene Citrate after 15 years: 27, 4 to 50).

#### Uncontrolled confounding

E-values suggested a strong unmeasured confounding variable (risk ratios > 1.50; up to five years post-treatment for time-varying models – see Table 3) would be required to explain the observed effects for uterine cancer at the lower-bound confidence intervals. Similarly strong unmeasured confounding variables would be needed to explain the associations for any ovarian cancer following ART or IUI/OS, serous ovarian cancer following ART, non-serous ovarian cancer following ART, thyroid cancer following Clomiphene Citrate, malignant melanoma following ART or IUI/OS, and in-situ melanoma following IUI/OS or Clomiphene Citrate. For most other cancers where a significant effect was observed, an unmeasured confounding variable of more modest strength (<1.50) would explain the observed effects at the lower bound confidence interval.

**Table 3:**
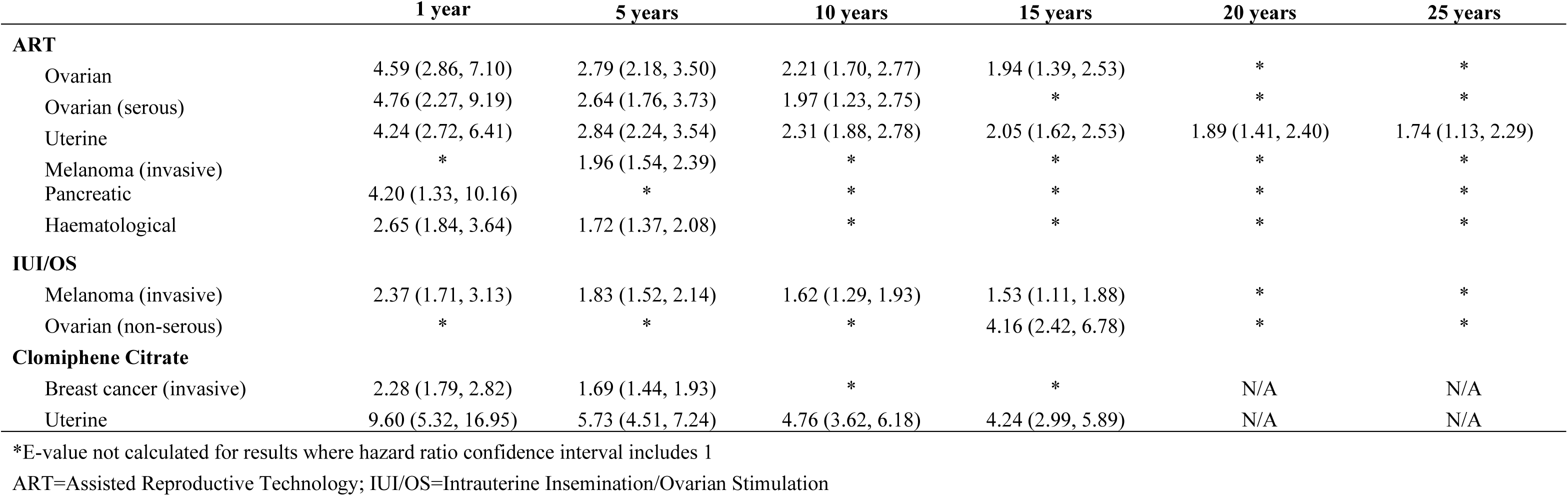
E-values over time for time-varying hazard ratio models.

|  | 1 year | 5 years | 10 years | 15 years | 20 years | 25 years |
| --- | --- | --- | --- | --- | --- | --- |
| **ART** |  |  |  |  |  |  |
| Ovarian | 4.59 (2.86, 7.10) | 2.79 (2.18, 3.50) | 2.21 (1.70, 2.77) | 1.94 (1.39, 2.53) | * | * |
| Ovarian (serous) | 4.76 (2.27, 9.19) | 2.64 (1.76, 3.73) | 1.97 (1.23, 2.75) | * | * | * |
| Uterine | 4.24 (2.72, 6.41) | 2.84 (2.24, 3.54) | 2.31 (1.88, 2.78) | 2.05 (1.62, 2.53) | 1.89 (1.41, 2.40) | 1.74 (1.13, 2.29) |
| Melanoma (invasive) | * | 1.96 (1.54, 2.39) | * | * | * | * |
| Pancreatic | 4.20 (1.33, 10.16) | * | * | * | * | * |
| Haematological | 2.65 (1.84, 3.64) | 1.72 (1.37, 2.08) | * | * | * | * |
| **IUI/OS** |  |  |  |  |  |  |
| Melanoma (invasive) | 2.37 (1.71, 3.13) | 1.83 (1.52, 2.14) | 1.62 (1.29, 1.93) | 1.53 (1.11, 1.88) | * | * |
| Ovarian (non-serous) | * | * | * | 4.16 (2.42, 6.78) | * | * |
| **Clomiphene Citrate** |  |  |  |  |  |  |
| Breast cancer (invasive) | 2.28 (1.79, 2.82) | 1.69 (1.44, 1.93) | * | * | N/A | N/A |
| Uterine | 9.60 (5.32, 16.95) | 5.73 (4.51, 7.24) | 4.76 (3.62, 6.18) | 4.24 (2.99, 5.89) | N/A | N/A |

\*E-value not calculated for results where hazard ratio confidence interval includes 1
ART=Assisted Reproductive Technology; IUI/OS=Intrauterine Insemination/Ovarian Stimulation

#### Sensitivity analysis

We conducted a sensitivity analysis removing all those who had a reportable cancer (both invasive and in-situ breast and melanoma) prior to t0. The results are reported in Supplementary Tables 9 to 11 and Supplementary Figure 1. No major differences were observed that would alter our conclusion. Small differences included that the model for invasive melanoma after ART was now best fit with no splines in the hazard ratio (but was still time varying), non-serous ovarian cancers after IUI/OS were now best fit by a non-time varying model, and invasive breast cancers following Clomiphene Citrate were now best fit by a model with no splines in the hazard radio (though still time varying). Cumulative marginal differences in expected cancers were similar (within 10% of the original estimate) across all models, even where the preferred model changed. The estimate for non-serous ovarian cancers following Clomiphene Citrate was no longer statistically significant.

## 6. DISCUSSION AND RECOMMENDATIONS

Using an emulated target trial design, we attempted to quantify the risk of hormone-related cancers after three types of MAR treatment (ART, IUI/OS, and Clomiphene Citrate). The excess risk observed for some cancers following treatment was notable, however E-values and analysis of non-hormone-related cancers suggested at least some of the excess risk is due to uncontrolled confounding and, for some cancers, bias due to differences in access to healthcare closer to the time of first treatment. Furthermore, our findings suggest that any excess risk of cancer following MAR treatments in real terms is relatively small, with most cancers showing evidence of fewer than 10 extra incident cases per 100,000 women per year, with only invasive breast cancer approaching nearly 20 extra cancers per 100,000 women per year. For comparison, this is similar to the estimated excess risk of breast cancer following regular use of combination and progesterone only hormonal contraceptives at ages 35-39.^34^

Analysis of unmeasured confounding through E-values suggests that some cancers would require strong residual confounding to account for the observed effects. On the other hand, evidence suggests conditions underlying infertility (including endometriosis and polycystic ovary syndrome (PCOS)) may be responsible for the observed associations between MAR treatments and uterine, ovarian, and thyroid cancers. Analysis of haematological cancers as a negative control suggests ethnicity may have confounded the association for melanoma, since both melanoma incidence^35^ and ART uptake^36^ differ across ethnic groups. As. Overall, it is likely we failed to establish a causal relationship using the emulated target trial framework (at least for melanoma, uterine, ovarian, and thyroid cancers) due to uncontrolled confounding.

However, we were able to leverage this framework to interrogate long-standing concerns with bias in studies on MAR and cancer. We also established that even with uncontrolled confounding, the absolute number of extra cancers expected following treatment is relatively small.

### Comparison with other studies

Recent meta-analyses suggest little to no effect of ART on hormone-related cancer incidence in women.^14, 15, 18^ Our finding that most cancers have at least a modest increase in incidence following MAR treatment conflicts somewhat with these results. Differences may be explained by the substantial heterogeneity in study designs and available confounding variables. This heterogeneity is compounded by changes in patterns of MAR use over time, between-country differences in eligibility for and access to MAR, and different approaches to the statistical analysis of observational datasets. In general, only more recent studies (approximately post 2010) have tried to adjust for relevant confounders like parity, mother’s age, and socioeconomic status.^15^ When meta-analysing only these studies, no significant effects of treatment on breast, ovarian, or uterine cancer were found.

If we consider only individual studies from the past ten years that have adjusted for confounding, we see some alignment to our findings. For example, Vassard et al.^37^ reported a significant relationship between ART and invasive breast cancer similar to that observed here, and Lundberg et al.^38^ found an even greater risk of ovarian cancer following ART in women who gave birth following treatment. Kessous et al.^39^ reported high excess risks for both ovarian and uterine cancer, but no increased risk for breast cancer (combining in-situ and invasive). Reigstad et al.^40^ found increased risk following ART treatment for thyroid cancer, but not other types of hormone-related cancer (noting wide confidence intervals).

They did however find a strong increased risk of uterine cancer following use of Clomiphene Citrate, an increased risk of invasive breast cancer following Clomiphene Citrate for parous women, and an increased risk for ovarian cancer for nulliparous women. Spaan et al.^41^ found evidence that borderline ovarian cancers, but not invasive ovarian cancers, are more common following ART. Overall, there is substantial variability in both confounder adjustment and analysis approach, with this variability contributing to heterogenous results.

### Cancer risk over time and detection bias

Understanding the pattern of cancer incidence over time since MAR is important, as there has long been concern the observed relationships between MAR and cancer may be biased by medical surveillance either while undergoing treatment or while pregnant^24, 25^ Using stratified Cox regression, Vassard et al.^24^ observed an increased risk of ovarian cancers immediately after ART treatment for women with endometriosis, followed by a gradual decrease in risk over time. They concluded increased surveillance was the most likely explanation. This observation is notable, as it is likely asymptomatic hormone-related cancers would have been identified in the work-up for ART or IUI/OS treatment.^25, 42, 43^ Similarly, in our study, several emulated trials suggested a greater likelihood of incident cancer shortly after first treatment. This pattern suggests two possibilities:

1. The treatment (alone or in combination with pregnancy) promotes the growth of existing but undetected cancers or transformation of benign to invasive cancers. That we observed an increased risk soon after some treatments for some cancers and not others potentially supports this hypothesis, as it suggests different treatments promote different types of cancer while being used. However, it is difficult to reconcile this hypothesis with similar time-varying effects for the non-hormone-related pancreatic and haematological cancers after MAR.
2. There is increased surveillance during treatment (and pregnancy), which leads to earlier detection of incident cancers (detection bias). In this case, we would observe a greater risk soon after treatment, and a lowering risk further from treatment due to depletion of cancers. That certain cancers were time-varying under certain treatments may be explained in this case by these cancers being more likely to be detected under more specialised medical surveillance (i.e., a fertility specialist when undergoing ART compared to a general practitioner when taking Clomiphene Citrate). This may contribute to the risk pattern for ovarian and uterine cancers in particular.

Overall, though it is possible that specific MAR treatments promote cancer growth, it is difficult to reconcile this explanation with our observed risk pattern for two of the three negative control cancers. Therefore, we believe that, like Vassard et al.^24^, detection bias is the more likely explanation for these results. This finding is the first empirical indication using a negative control that medical surveillance may be responsible for an increased risk of cancer following MAR.

### Unmeasured confounding

Our E-value analysis suggested some of the associations we observed could be explained by uncontrolled confounding. One such factor is the underlying cause of infertility, which was not recorded in any of the available datasets. A recent meta-analysis suggested endometriosis has a sufficiently strong relationship to both any ovarian (risk ratio 1.93 – noting non-serous ovarian cancer after ART had a lower-bound E-value of 1.94) and thyroid cancer (1.39) to explain our observed results for these cancers following ART and IUI/OS.^44, 45^ The risk ratio for infertility for women with endometriosis compared to the general population has been estimated at 2.12,^46^ indicating for these cancers both the confounder-exposure and confounder-outcome relationship may have sufficient strength to account for the associations we observed.

PCOS may also contribute to unmeasured confounding, particularly for uterine cancers and for Clomiphene Citrate users. Though it is uncommon to use Clomiphene Citrate for those with endometriosis-induced infertility (as endometriosis rarely causes anovulation), PCOS is partly defined by the presence of irregular ovulation,^47^ and Clomiphene Citrate is commonly prescribed for those with PCOS-induced infertility. A recent meta-analysis found PCOS was strongly associated with risk of uterine cancer (odds ratio 4.07).^48^ Furthermore, anovulation is a strong risk factor for uterine cancer^49^ and more common in obese women and women with diabetes,^50, 51^ who are at greater risk of uterine cancer^52^ and thyroid cancer.^53, 54^ Finally, a 2019 registry study found PCOS was significantly associated with ovarian cancer (hazard ratio 2.16), though not thyroid cancer.^55^ Overall, underlying causes of infertility, particularly endometriosis, PCOS, coupled with associated obesity and diabetes, may account for either all or a substantial proportion of the observed effect of MAR treatment on the development of ovarian, uterine, and thyroid cancers.

Our findings for breast cancer (invasive and in-situ), colorectal, and melanoma (invasive and in-situ) may be more robust to unmeasured confounding, as there is less evidence the underlying cause of infertility is associated with these cancers. Some recent studies have found no relationship between infertility history and overall risk of breast cancer^56^ or colorectal cancer.^57^ Notably, in our study both these cancers had the lowest observed increase in relative risk following the MAR treatments. For IUI/OS, there was no effect of treatment on the incidence of colorectal cancer, and for invasive breast cancer following Clomiphene Citrate, the increased risk disappeared over time. Due to the high background rate of invasive breast cancer in the population, this small increase in relative risk translated to around an average of 400 extra cancers per 100,000 women treated over the course of the study period. Findings on melanoma and its associations with fertility and pregnancy hormones (like oestrogen) are mixed,^58^ so it is difficult to determine whether there is infertility related confounding for this malignancy.

Haematological cancer showed a positive association with MAR treatment, despite no clear theorised underlying biological mechanisms. That is, even if the underlying cause of infertility may not confound the relationship between MAR and breast, colorectal, and melanoma, it may be other sources of bias we have not accounted for which may drive these observed associations. International estimates show Caucasian women are more likely to undergo fertility treatments.^36^ As melanoma and leukemia, Hodgkin lymphoma and non-Hodgkin lymphoma are more common in White or Caucasian people,^35, 59–61^ confounding by ethnicity may be responsible for the associations we observed for these malignancies.

Finally, Australian women are invited to attend breast and colorectal cancer screening and there is opportunistic skin cancer screening for individuals at high risk. According to Andersen’s behavioural model for use of health services,^62^ it is likely women who have engaged in fertility treatments are more likely to engage with these screening programs, and this disparity in seeking health services may also have affected our findings.

We note that a negative association between lung cancer and MAR treatment was observed. Though it is possible that uncontrolled confounding was responsible for this association, we posit a more likely explanation is that women undergoing fertility treatments were required to stop smoking prior to treatment to improve the both the probability of conception and health of the foetus on the advice of their healthcare professional. This cessation still represents an effect of treatment, but not due to the hormone drugs used in the treatment under scrutiny.

### Strengths and limitations of study

We used population-based data linkage to identify the exposures, outcomes and covariates, capturing over 25 years of data. However, we were limited to routinely collected national data, which did not include several important confounders such as underlying cause of infertility, ethnicity, individual-level socioeconomic factors (e.g., education and income, which may also be markers for engagement with preventive health services), family history of cancer, smoking, and oral contraceptive use. Although the optimal emulated trial would account for all potential confounding factors, this is likely an impossible task for causal questions regarding use of MAR.^12^ For example, it is estimated 40% of suspected endometriosis cases are not captured in administrative medical records.^63^ Due to concerns of unmeasured confounding from cause of infertility, as well as computational feasibility, we were unable to assess a mediating effect of pregnancy. Given pregnancy is thought to be associated with a changed risk of several hormone-related cancers^64^ it is likely there may be a different effect of MAR treatment on cancer dependent on whether a pregnancy is achieved. Finally, our study was conducted within an Australian context, which subsidises the use of most MAR treatments with no exceptions for age or prior children conceived, but historically did not subsidise those not deemed to be medically infertile. Thus, caution must be taken when applying the results presented here directly to other countries with different MAR eligibility criteria.

### Strengths and weaknesses in relation to other studies

To our knowledge, this study represents the first attempt to explicitly model hazard over time, calculate the cumulative marginal expected number of cancers for MAR-exposed women, calculate E-values to estimate unmeasured confounding, and apply the target trial emulation framework to the question of cancer risk following MAR. However, we could not access underlying cause of infertility, which is at least partially available in some other national datasets being used to examine the association between MAR and cancer risk (e.g., CoNARTaS^65^; OMEGA^66^). Furthermore, because our data were limited to fertility-related treatments, we could not ascertain surgical history for hysterectomy, oophorectomy, or mastectomy, which would preclude the development of the relevant cancers. Finally, we only included women who received government reimbursement for their MAR treatments. Though this is the vast majority of women undergoing MAR treatment, we will have misclassified some women who paid out-of-pocket for fertility services (such as those in same-sex relationships without a diagnosis of infertility, which is not covered under Medicare), and this will have biased our findings towards the null.

### Meaning of the study

This research has important implications for clinical practice, public health policy, and future research. Clinically, the findings highlight the increased cancer risks observed following MAR treatments may be partially or fully attributed to underlying infertility conditions (ovarian, uterine and thyroid cancer), heightened medical surveillance (breast and colorectal cancer), and demographic characteristics (melanoma), rather than MAR itself. Health care professionals should therefore emphasize cautious interpretation and communication of cancer risk relative to women who consider undertaking MAR. Furthermore, there is a clear need for ongoing surveillance of women after MAR treatments.

### Unanswered questions and future research

Our study highlights two areas for further research. It remains unclear whether there may still be an effect of MAR treatment on ovarian, uterine and thyroid cancers (albeit smaller than we observed). To improve our ability to answer this question, it is critical that we collect and make available for research information on key confounding variables such as fertility treatment indications, obesity, use of hormonal contraceptives, and demographic and lifestyle factors. Such enhanced data collection efforts would significantly strengthen the capacity to identify and account for complex confounding pathways, ultimately improving the validity of causal inferences related to MAR and risk of cancer and other health outcomes. Second, the small effect sizes for breast and colorectal cancer mean these findings could be particularly vulnerable to uncontrolled confounding. Finally, future studies should aim to replicate the time-varying effects across cancers to test for robustness.

### Conclusions

Our study provides the first attempt to use the emulated target trial framework to quantify the effect of three different MAR treatments on risk of hormone-related cancers in women. Despite observing increased relative risk for most hormone-related cancers following MAR, this corresponded to only small increases in absolute excess risk. Bias analysis and the inclusion of negative controls showed the elevated risk was potentially fully attributable to uncontrolled confounding in some cases. Clinicians should be aware that a small excess risk of cancer following MAR treatment may exist, and this excess risk cannot be confidently attributed to MAR.

## Supporting information

ICMJE Declaration forms

## 7. AUTHOR CONTRIBUTIONS

ARW was involved in data curation, conceptualisation, software, visualisation, and formal analyses and wrote the first draft of the manuscript. CMV, GMC, CV, SO, AA, MC, NH, and LJ were involved in study conceptualisation and funding acquisition.

CMV and GMC were involved in resources.

CMV, GMC, CV and SO were involved in supervision and interpretation.

All authors critically reviewed and edited the manuscript, approved the final version for submission, and agreed to be accountable for all aspects of this work.

ARW is the guarantor.

The corresponding author attests that all listed authors meet authorship criteria and that no others meeting the criteria have been omitted.

## Data Availability

The analyses were based on data from different registries and administrative datasets (Australian Department of Health and Aged Care, State and Territory Health Departments, and the Australian Institute of Health and Welfare). And the data can be made available on request to each of the data custodians after ethical approval from the relevant Human Research Ethics Committees. The statistical analysis plan is available at https://osf.io/rk9n6, and analysis code at https://osf.io/nqkc2/.

## ACKNOWLEDGEMENTS

The authors are grateful for the perspectives and advice offered by Maree Pickens, consumer advocate, in the interpretation of the findings. We also acknowledge and thank the staff at the Australian Institute of Health and Welfare Data Linkage Unit, the Statistical Analysis and Linkage Unit of the Statistical Services Branch, Queensland Health, the NSW Centre for Health Record Linkage, the Tasmanian Data Linkage Unit, the Centre for Victorian Data Linkage, and the Linkage, Data Engineering Outputs and ISPD Client Services teams at Western Australia Data Linkage Services for supporting the project and undertaking data linkage. We thank the Australian Institute of Health and Welfare and the population-based cancer registries of New South Wales (NSW), Victoria, Queensland, Western Australia, South Australia, Tasmania, the Australian Capital Territory (ACT) and the Northern Territory for the provision of data from the Australian Cancer Database. We also thank the following data custodians for providing the datasets used for this project: Australian Department of Health and Aged Care (Medicare Benefits Schedule, Pharmaceutical Benefits Scheme); Australian Institute of Health and Welfare (National Death Index); the NSW Ministry of Health, the Department of Health Victoria, the Statistical Services Branch Queensland Health, the Department of Health Western Australia, Preventive Health South Australia, the Department of Health Tasmania, HealthInfo ACT (perinatal data); and the jurisdictional Registries of Births, Deaths and Marriages (birth data). We are grateful to the Victorian Consultative Council on Obstetric and Paediatric Mortality and Morbidity (CCOPMM) for providing access to the data used for this project and for the assistance of the staff at Safer Care Victoria. The conclusions, findings, opinions and views or recommendations expressed in this paper are strictly those of the authors. They do not necessarily reflect those of CCOPMM.

## 11. DISSEMINATION TO PARTICIPANTS AND RELATED PATIENT AND PUBLIC COMMUNICITIES

This manuscript will be made available through the author’s institutions. The authorship team will work with consumer advocates to disseminate the findings including via press release and a lay summary.

## 12. TRANSPARENCY

ARW and CV declare that the manuscript is an honest, accurate, and transparent account of the study being reported, no important aspects of the study have been omitted, and any discrepancies from the original study plan have been explained. Enquiries about methodology should be directed to.

## 13. SUPPLEMENTARY METHODS

### Supplementary Methods 1: Flexible parametric survival models

We had originally planned to use Cox regression in our analysis. However, there were clear benefits to switching to flexible parametric survival models. Flexible parametric survival models estimate a parametric model of the baseline hazard, which allows for prediction of survival over time. These models also allow underlying hazards to vary for specified categorical variables, which allows the hazard ratio to vary over time. When no variables are specified as having time-varying hazards, flexible parametric models closely approximate Cox regression. Our model of the baseline hazard included two splines. We also included two splines for the hazard function of age at first treatment based on preliminary modelling suggesting that this would provide a better model fit. We tested four models to ascertain the number of splines needed in the hazard function of treatment to determine the best fitting model. The models tested include a model with no time-varying effect of treatment, and three models with time varying effects of treatment, altering the number of splines in the hazard function (0, 1, or 3 splines, equally spaced). Flexible parametric models were calculated using the *stpm3* command in STATA.

The model selection steps included:

1. Comparing each time-varying model to the non-time-varying model. If one model outperformed the non-time-varying model, we selected that model. If multiple models outperformed the non-time-varying model, we went to step 2. Otherwise, we selected the non-time-varying model;
2. Comparing the remaining models to the simplest model that outperformed the non-time varying model. If one model outperformed the simplest model, we selected that model. If multiple models outperformed the simplest model, we repeated step 2. Otherwise, we selected the simplest model.

### Supplementary Methods 2: Changes from registered protocol

We note that, compared to our registered protocol, we opted to use flexible parametric survival models over Cox regression to allow the recovery of cumulative marginal differences, and a changing proportional hazard over time. We were unable to assess the effect of gonadotrophins or anti-gonadotropin-releasing hormone use due to issues with completeness of data capture for these medicines. We chose not to assess dose effect of treatment due to the computational feasibility of conducting a g-methods-based analysis with large data across a long period of time for multiple models, and the impact of unmeasured confounding from cause of infertility (which would likely be exacerbated in the calculation of a dose effect). People with cancers of the same topography as the target cancer before t0 were not excluded, as our cancer definitions included multiple histologies within the same topography that could be considered primary cancers. Sensitivity analyses on limiting the time of follow-up to ascertain changes in the hazard ratio were not run due to flexible parametric survival models accounting for the changing hazard ratios over time. The sensitivity analysis excluding those indicated as having poor linkage to children’s birth records was not run as a linkage quality indicator was not provided to researchers, though researchers removed women who had a birth recorded after their death was recorded prior to analysis.

## 14. SUPPLEMENTARY TABLES

**Supplementary Table 1:**
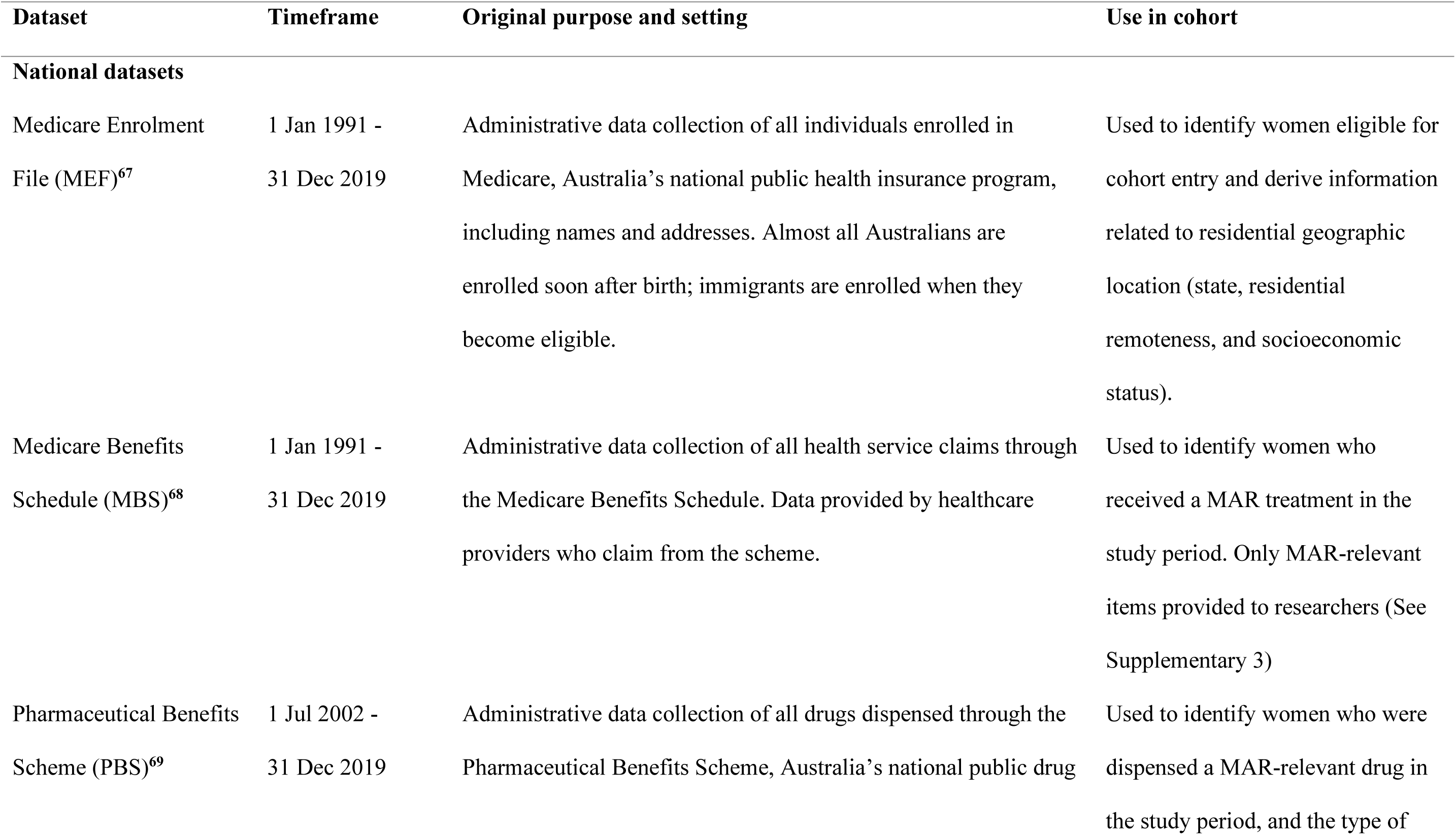
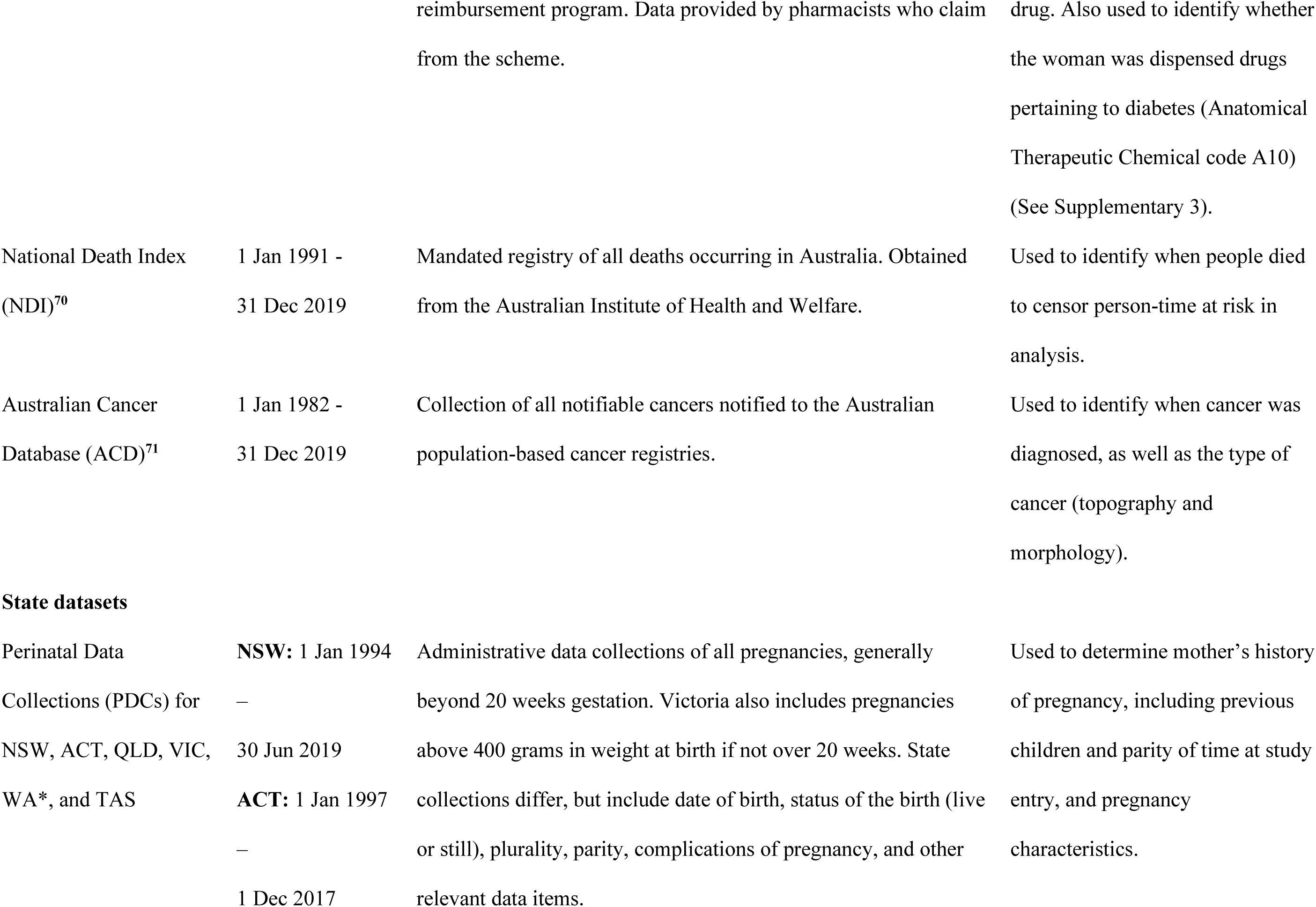
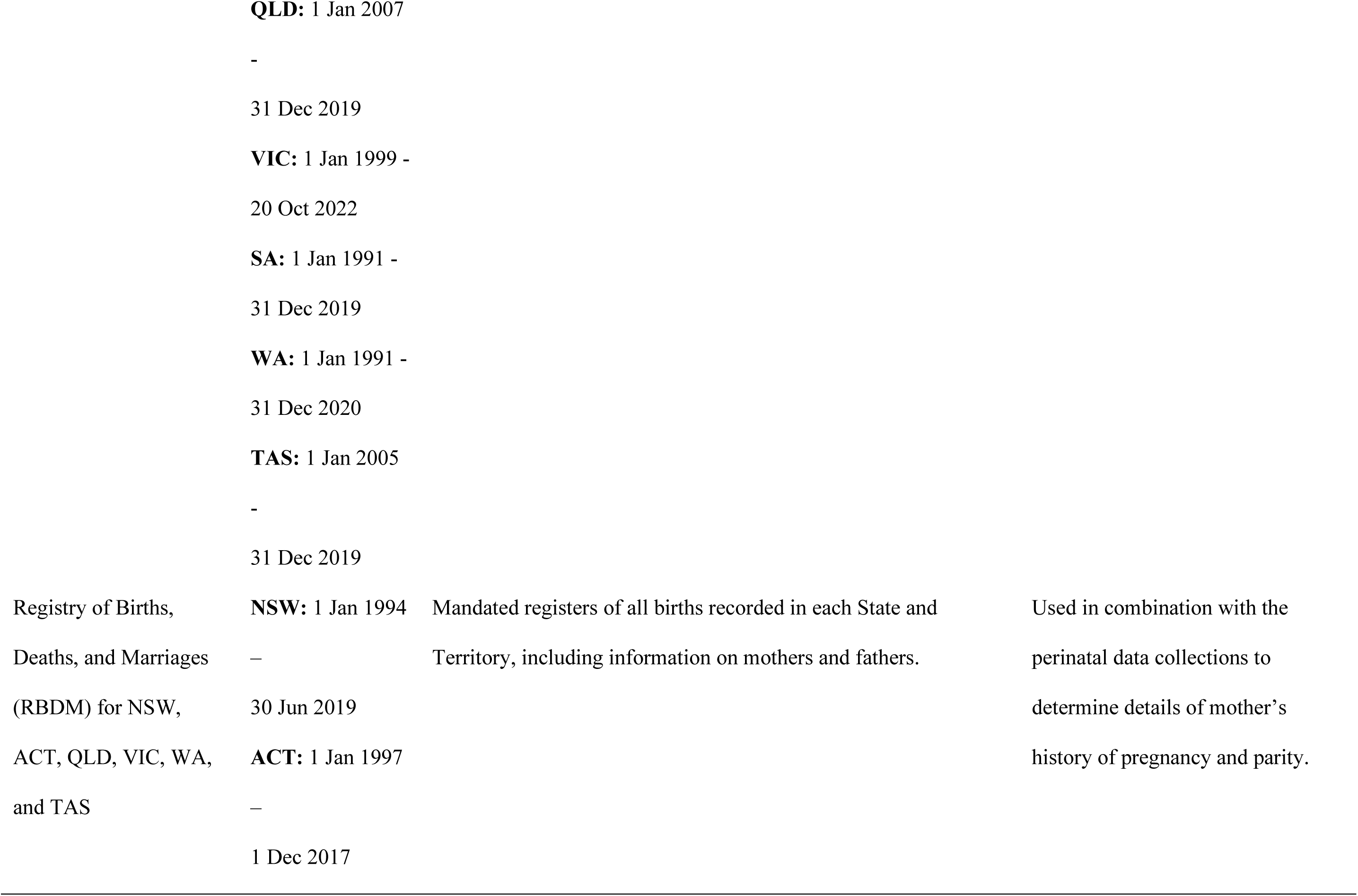
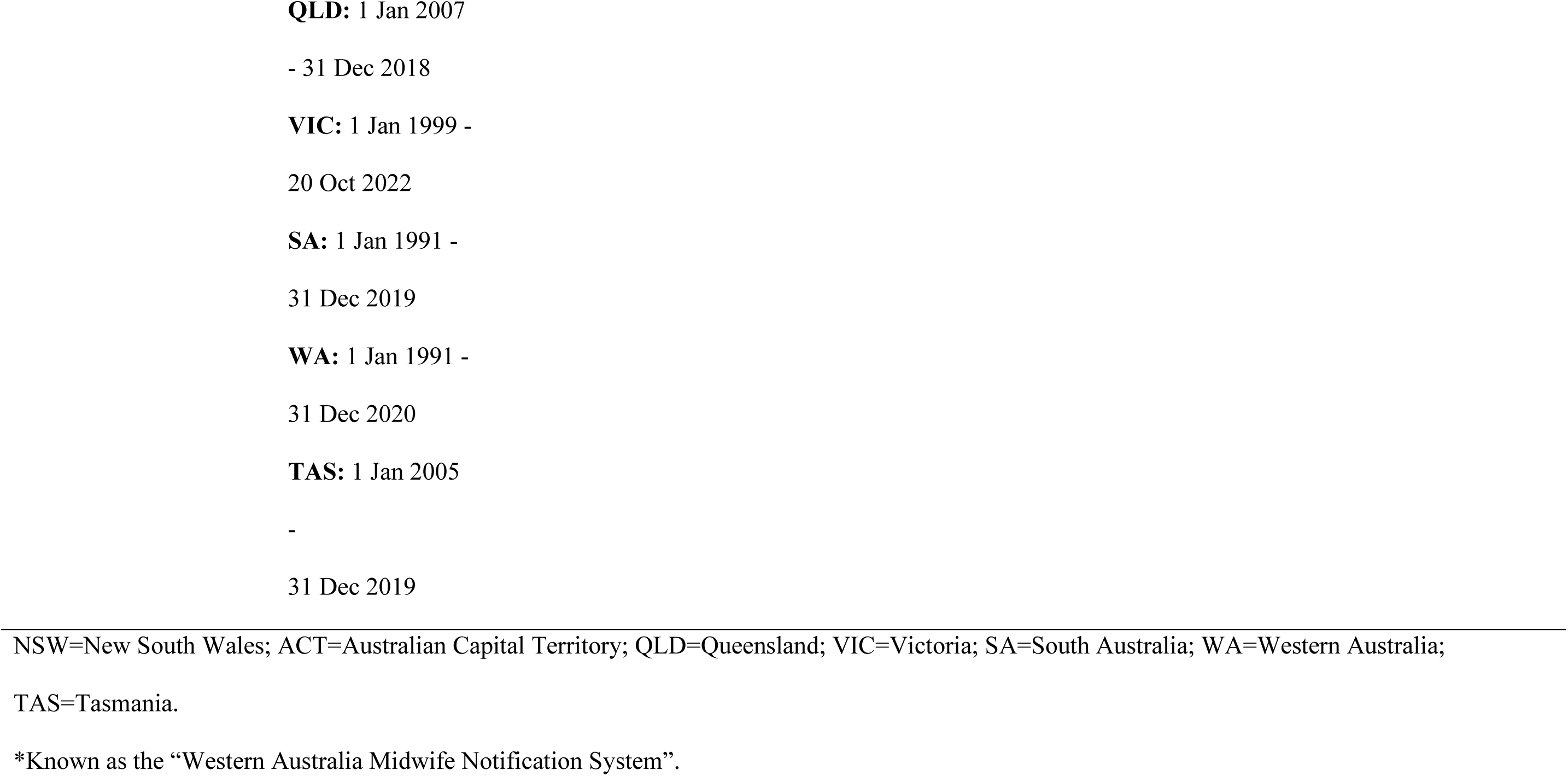
Description of datasets used in linkage.

**Supplementary Table 2:** Target trial specification.

|  | **Target trial specification** |
| --- | --- |
| **Eligibility criteria** | Australian women aged between 18 and 55 between January 1991 and 31 December 2018 who were eligible to receive a MAR treatment and had not previously received a MAR treatment. |
| **Treatment strategies** | Treatment group: undergoing a MAR treatment (ART, IUI/OS) or receiving a Clomiphene prescription, depending on the type of treatment under examination;<br>Comparator group: not undergoing a MAR treatment of any type. |
| **Assignment procedures** | Participants are randomly assigned to the treatment group and comparator group (four comparator participants assigned for every treatment participant). |
| **Follow-up** | Until end of study period or death. |
| **Outcomes** | Diagnosis of the target cancer (invasive breast, in-situ breast, any ovarian, serous ovarian, non-serous ovarian, uterine, thyroid, colorectal, invasive melanoma, in-situ melanoma). |
| **Causal contrasts** | Per-protocol |
| **Identifying assumptions** | Randomisation occurred during group assignment, and censoring events during follow-up do not occur systematically based on the presence or absence of the exposure. |
| **Data analysis plan** | Appropriate survival analysis that estimates both the hazard ratio and survival difference of those assigned to the treatment group compared to the comparator group. |

MAR=Medically Assisted Reproduction; ART=Assisted Reproductive Technology; IUI/OS=Intrauterine Insemination/Ovarian Stimulation

**Supplementary Table 3:**
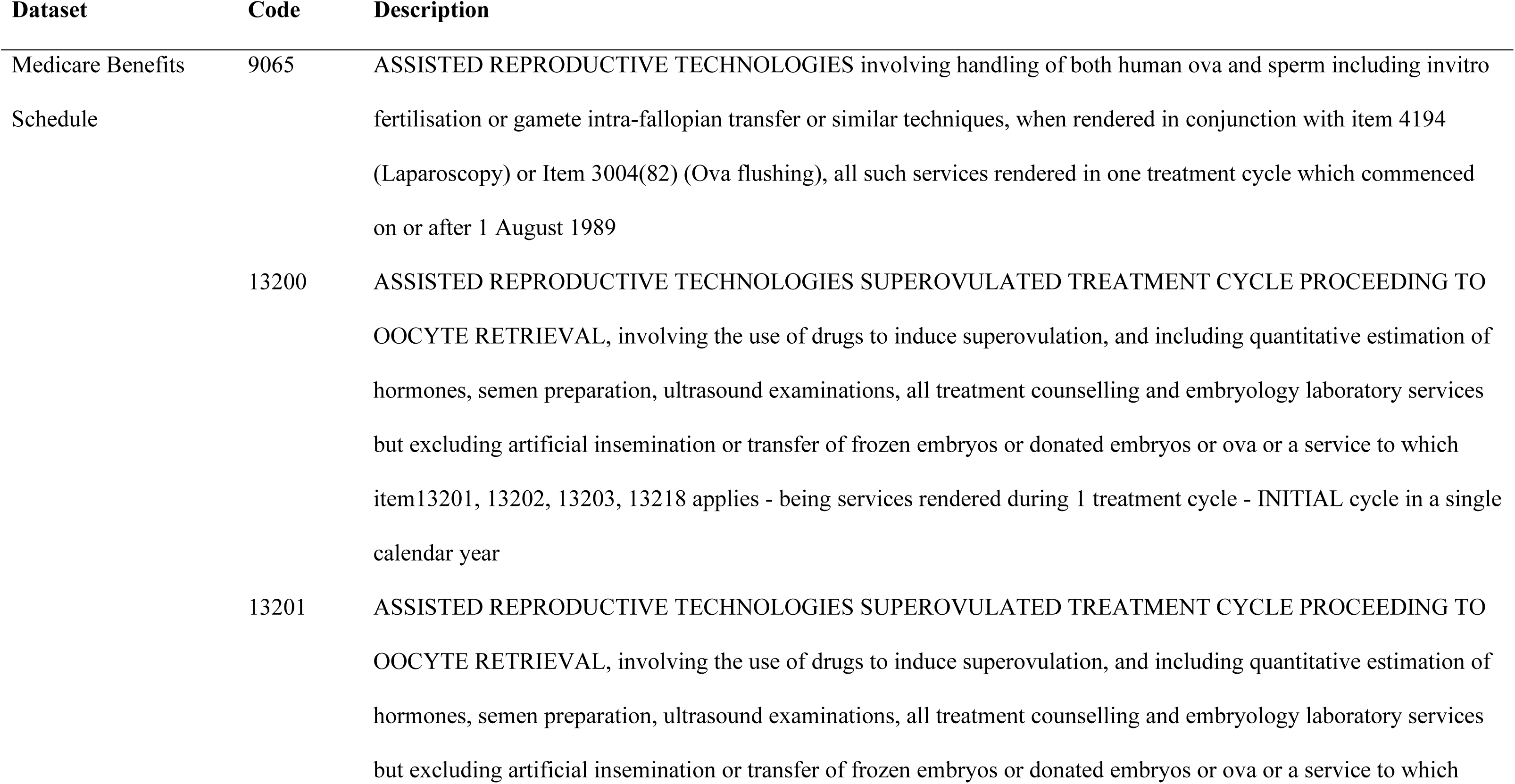
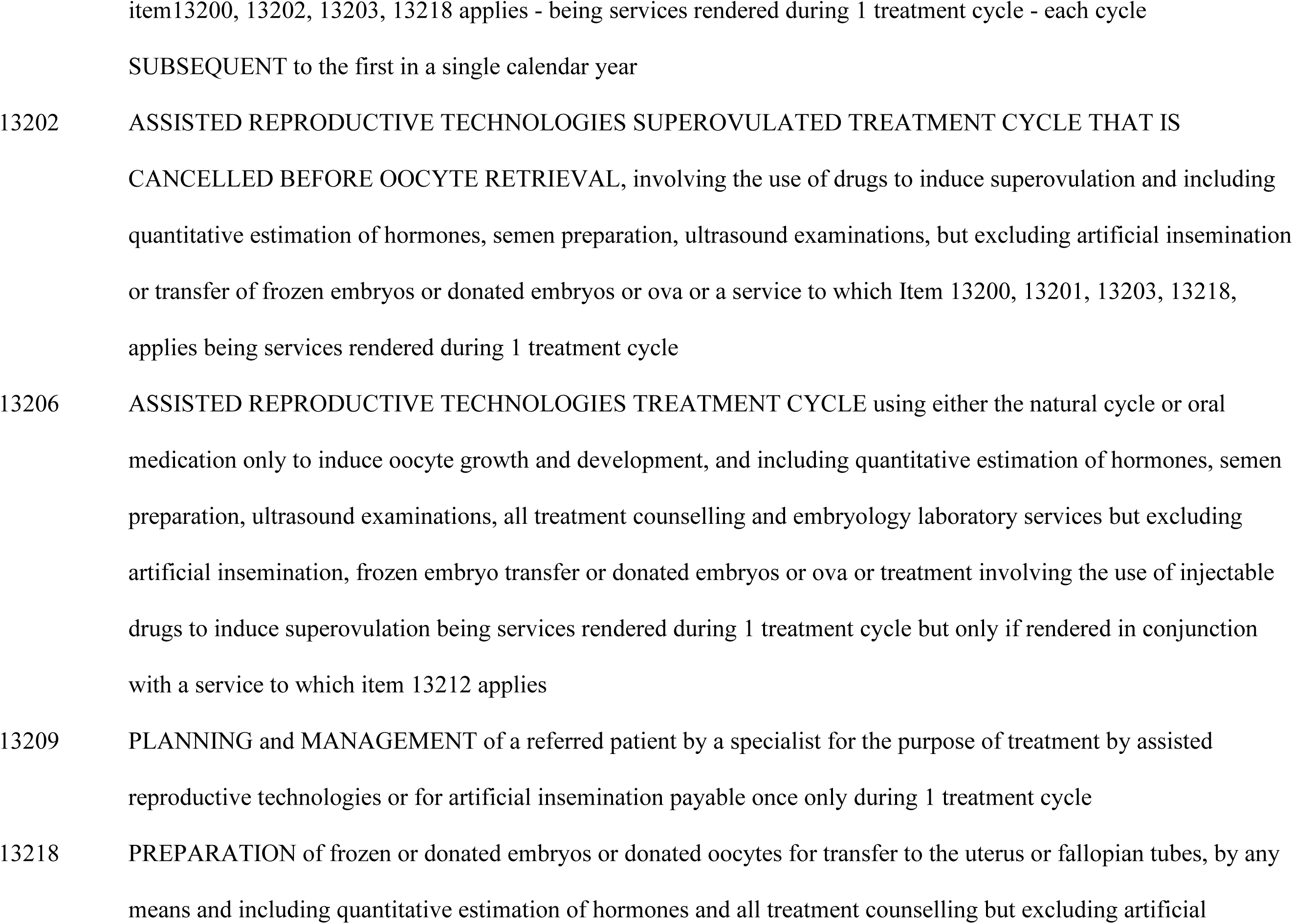
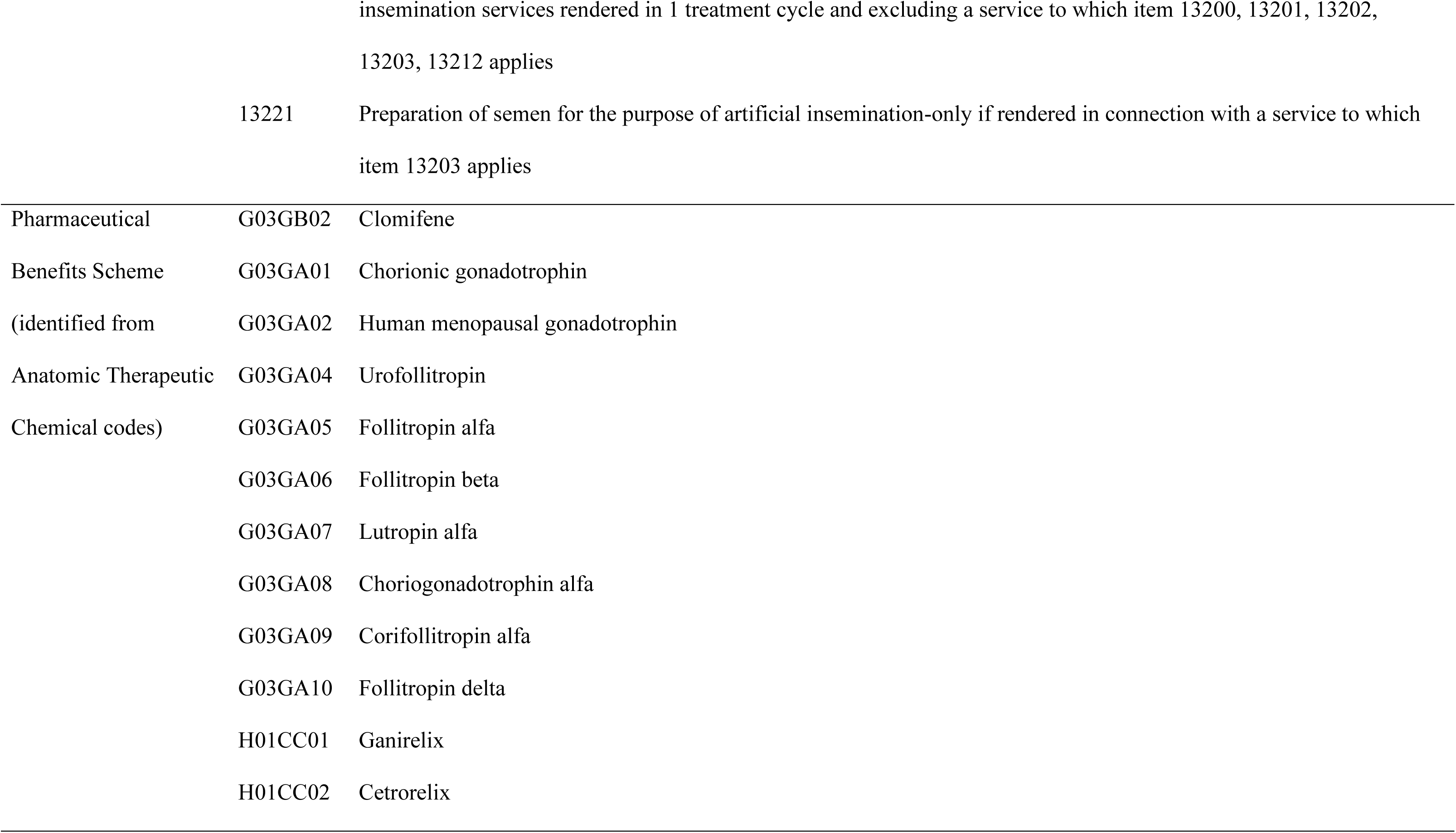
Medicare Benefits Schedule (MBS) codes and Anatomic Therapeutic Chemical (ATC) codes used to identify women exposed medically assisted reproduction.

**Supplementary Table 4:** Medicare benefits schedule (MBS) and Pharmaceutical Benefits Scheme)

|  | MBS item codes | PBS item codes |
| --- | --- | --- |
| **Before Jan 2010** |  |  |
| ART (IVF + ICSI) | 13200 OR |  |
|  | 13206 OR |  |
|  | 13212 |  |
| IUI/OS | 13203 |  |
| Clomiphene Citrate use |  | 01211R |
| **After Jan 2010** |  |  |
| ART (IVF + ICSI) | 13200 OR |  |
|  | 13201 OR |  |
|  | 13206 OR |  |
|  | 13212 |  |
| IUI/OS | 13202 OR |  |
|  | 13203 |  |
| Clomiphene Citrate use |  | 01211R |

MBS=Medicare Benefits Schedule; PBS=Pharmaceutical Benefits Scheme; MAR=Medically assisted reproduction; ART=Assisted reproduction technology; IVF=In-vitro fertilisation; IUI/OS=Intrauterine Insemination/Ovarian Stimulation

**Supplementary Table 5:** International classification of Diseases – Oncology 3^rd^ edition topographies and morphologies for cancer classification.

| **Target cancer** | **ICD-O-3 topography** | **ICD-O-3.1 morphology (all behaviour code 3 unless otherwise stated)** |
| --- | --- | --- |
| **Breast (invasive)** | C50 | All |
| **Breast (in-situ)** | C50 | All (behaviour code 2) |
| **Ovarian (all)** | C56 | All |
| **Ovarian (serous)** | C56 | 8441 |
| **Ovarian (non-serous)** | C56 | All except 8441 |
| **Uterine** | C54, C55 | All |
| **Melanoma (invasive)** | C44 | 872-879 |
| **Melanoma (in-situ)** | C44 | 872-879 (behaviour code 2) |
| **Colorectal (excluding anus)** | C18-C20 | All |
| **Thyroid** | C73 | All |
| **Pancreatic** | C25 | All |
| **Lung** | C33-C34 | All |
| **Haematological** | C00-C80 | 959-998 |

**Supplementary Table 6:** Numbers of incident cancers after study entry. Percentages are relative to the size of the respective treatment/comparator t.

| Cancer type | ART |  | IUI/OS |  | Clomiphene Citrate |  |
| --- | --- | --- | --- | --- | --- | --- |
|  | Treatment | Comparator | Treatment | Comparator | Treatment | Comparator |
| Breast (invasive) | 2,588 (1.5%) | 7,541 (1.3%) | 1,445 (1.9%) | 4,245 (1.6%) | 1,191 (0.8%) | 3,847 (0.7%) |
| Breast (in-situ) | 212 (0.2%) | 567 (0.1%) | 79 (0.2%) | 230 (0.1%) | 170 (0.1%) | 529 (0.1%) |
| Ovarian | 221 (0.1%) | 461 (0.1%) | 124 (0.2%) | 266 (0.1%) | 82 (0.1%) | 198 (0.0%) |
| Ovarian (serous) | 103 (0.1%) | 237 (0.0%) | 60 (0.1%) | 127 (0.0%) | 32 (0.0%) | 81 (0.0%) |
| Ovarian (non-serous) | 118 (0.1%) | 225 (0.0%) | 64 (0.1%) | 139 (0.1%) | 50 (0.0%) | 117 (0.0%) |
| Uterine | 297 (0.2%) | 632 (0.1%) | 180 (0.2%) | 364 (0.1%) | 193 (0.1%) | 192 (0.0%) |
| Thyroid | 466 (0.3%) | 1,295 (0.2%) | 232 (0.3%) | 695 (0.3%) | 362 (0.2%) | 977 (0.2%) |
| Colorectal (excluding anus) | 447 (0.3%) | 1,288 (0.2%) | 234 (0.3%) | 758 (0.3%) | 228 (0.1%) | 661 (0.1%) |
| Melanoma (invasive) | 871 (0.5%) | 2,546 (0.4%) | 538 (0.7%) | 1,437 (0.6%) | 565 (0.4%) | 1,618 (0.3%) |
| Melanoma (in-situ) | 485 (0.4%) | 1,490 (0.3%) | 217 (0.5%) | 530 (0.3%) | 580 (0.4%) | 1,592 (0.3%) |
| Pancreatic | 51 (0.0%) | 186 (0.0%) | 32 (0.0%) | 120 (0.0%) | 20 (0.0%) | 69 (0.0%) |
| Lung | 173 (0.1%) | 684 (0.1%) | 90 (0.1%) | 401 (0.2%) | 64 (0.0%) | 231 (0.0%) |
| Haematological | 465 (0.3%) | 1,297 (0.2%) | 286 (0.4%) | 716 (0.3%) | 257 (0.2%) | 739 (0.1%) |

ART=Assisted Reproductive Technology; IUI/OS=Intrauterine Insemination/Ovarian Stimulation

**Supplementary Table 7:**
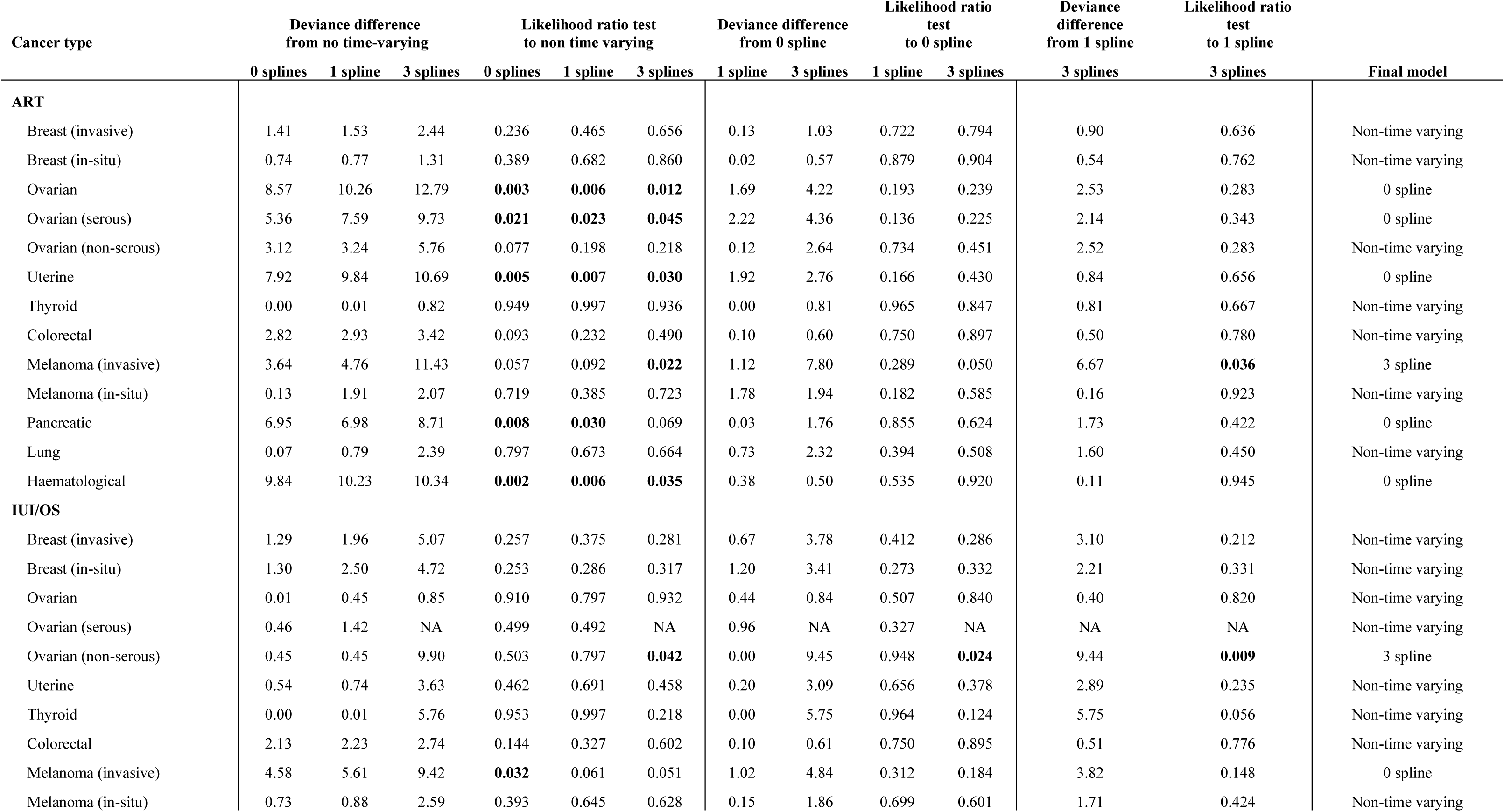
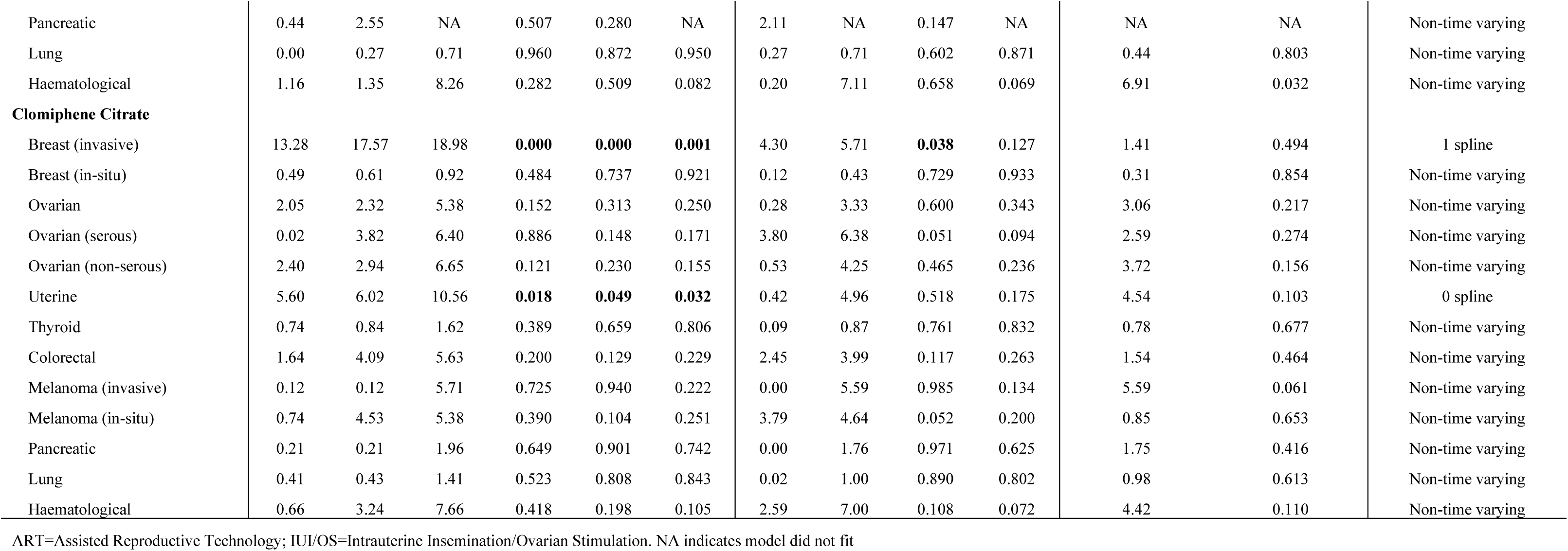
Deviance and likelihood ratio tests for selection of models.

| Cancer type | Deviance difference<br>from no time-varying |  |  | Likelihood ratio test<br>to non time varying |  |  | Deviance difference<br>from 0 spline |  | Likelihood ratio<br>test<br>to 0 spline |  | Deviance<br>difference<br>from 1 spline | Likelihood ratio<br>test<br>to 1 spline | Final model |
| --- | --- | --- | --- | --- | --- | --- | --- | --- | --- | --- | --- | --- | --- |
|  | 0 splines | 1 spline | 3 splines | 0 splines | 1 spline | 3 splines | 1 spline | 3 splines | 1 spline | 3 splines | 3 splines | 3 splines |  |
| ART |  |  |  |  |  |  |  |  |  |  |  |  |  |
| Breast (invasive) | 1.41 | 1.53 | 2.44 | 0.236 | 0.465 | 0.656 | 0.13 | 1.03 | 0.722 | 0.794 | 0.90 | 0.636 | Non-time varying |
| Breast (in-situ) | 0.74 | 0.77 | 1.31 | 0.389 | 0.682 | 0.860 | 0.02 | 0.57 | 0.879 | 0.904 | 0.54 | 0.762 | Non-time varying |
| Ovarian | 8.57 | 10.26 | 12.79 | **0.003** | **0.006** | **0.012** | 1.69 | 4.22 | 0.193 | 0.239 | 2.53 | 0.283 | 0 spline |
| Ovarian (serous) | 5.36 | 7.59 | 9.73 | **0.021** | **0.023** | **0.045** | 2.22 | 4.36 | 0.136 | 0.225 | 2.14 | 0.343 | 0 spline |
| Ovarian (non-serous) | 3.12 | 3.24 | 5.76 | 0.077 | 0.198 | 0.218 | 0.12 | 2.64 | 0.734 | 0.451 | 2.52 | 0.283 | Non-time varying |
| Uterine | 7.92 | 9.84 | 10.69 | **0.005** | **0.007** | **0.030** | 1.92 | 2.76 | 0.166 | 0.430 | 0.84 | 0.656 | 0 spline |
| Thyroid | 0.00 | 0.01 | 0.82 | 0.949 | 0.997 | 0.936 | 0.00 | 0.81 | 0.965 | 0.847 | 0.81 | 0.667 | Non-time varying |
| Colorectal | 2.82 | 2.93 | 3.42 | 0.093 | 0.232 | 0.490 | 0.10 | 0.60 | 0.750 | 0.897 | 0.50 | 0.780 | Non-time varying |
| Melanoma (invasive) | 3.64 | 4.76 | 11.43 | 0.057 | 0.092 | **0.022** | 1.12 | 7.80 | 0.289 | 0.050 | 6.67 | **0.036** | 3 spline |
| Melanoma (in-situ) | 0.13 | 1.91 | 2.07 | 0.719 | 0.385 | 0.723 | 1.78 | 1.94 | 0.182 | 0.585 | 0.16 | 0.923 | Non-time varying |
| Pancreatic | 6.95 | 6.98 | 8.71 | **0.008** | **0.030** | 0.069 | 0.03 | 1.76 | 0.855 | 0.624 | 1.73 | 0.422 | 0 spline |
| Lung | 0.07 | 0.79 | 2.39 | 0.797 | 0.673 | 0.664 | 0.73 | 2.32 | 0.394 | 0.508 | 1.60 | 0.450 | Non-time varying |
| Haematological | 9.84 | 10.23 | 10.34 | **0.002** | **0.006** | **0.035** | 0.38 | 0.50 | 0.535 | 0.920 | 0.11 | 0.945 | 0 spline |
| IUI/OS |  |  |  |  |  |  |  |  |  |  |  |  |  |
| Breast (invasive) | 1.29 | 1.96 | 5.07 | 0.257 | 0.375 | 0.281 | 0.67 | 3.78 | 0.412 | 0.286 | 3.10 | 0.212 | Non-time varying |
| Breast (in-situ) | 1.30 | 2.50 | 4.72 | 0.253 | 0.286 | 0.317 | 1.20 | 3.41 | 0.273 | 0.332 | 2.21 | 0.331 | Non-time varying |
| Ovarian | 0.01 | 0.45 | 0.85 | 0.910 | 0.797 | 0.932 | 0.44 | 0.84 | 0.507 | 0.840 | 0.40 | 0.820 | Non-time varying |
| Ovarian (serous) | 0.46 | 1.42 | NA | 0.499 | 0.492 | NA | 0.96 | NA | 0.327 | NA | NA | NA | Non-time varying |
| Ovarian (non-serous) | 0.45 | 0.45 | 9.90 | 0.503 | 0.797 | **0.042** | 0.00 | 9.45 | 0.948 | **0.024** | 9.44 | **0.009** | 3 spline |
| Uterine | 0.54 | 0.74 | 3.63 | 0.462 | 0.691 | 0.458 | 0.20 | 3.09 | 0.656 | 0.378 | 2.89 | 0.235 | Non-time varying |
| Thyroid | 0.00 | 0.01 | 5.76 | 0.953 | 0.997 | 0.218 | 0.00 | 5.75 | 0.964 | 0.124 | 5.75 | 0.056 | Non-time varying |
| Colorectal | 2.13 | 2.23 | 2.74 | 0.144 | 0.327 | 0.602 | 0.10 | 0.61 | 0.750 | 0.895 | 0.51 | 0.776 | Non-time varying |
| Melanoma (invasive) | 4.58 | 5.61 | 9.42 | **0.032** | 0.061 | 0.051 | 1.02 | 4.84 | 0.312 | 0.184 | 3.82 | 0.148 | 0 spline |
| Melanoma (in-situ) | 0.73 | 0.88 | 2.59 | 0.393 | 0.645 | 0.628 | 0.15 | 1.86 | 0.699 | 0.601 | 1.71 | 0.424 | Non-time varying |
| Pancreatic | 0.44 | 2.55 | NA | 0.507 | 0.280 | NA | 2.11 | NA | 0.147 | NA | NA | NA | Non-time varying |
| Lung | 0.00 | 0.27 | 0.71 | 0.960 | 0.872 | 0.950 | 0.27 | 0.71 | 0.602 | 0.871 | 0.44 | 0.803 | Non-time varying |
| Haematological | 1.16 | 1.35 | 8.26 | 0.282 | 0.509 | 0.082 | 0.20 | 7.11 | 0.658 | 0.069 | 6.91 | 0.032 | Non-time varying |
| **Clomiphene Citrate** |  |  |  |  |  |  |  |  |  |  |  |  |  |
| Breast (invasive) | 13.28 | 17.57 | 18.98 | **0.000** | **0.000** | **0.001** | 4.30 | 5.71 | **0.038** | 0.127 | 1.41 | 0.494 | 1 spline |
| Breast (in-situ) | 0.49 | 0.61 | 0.92 | 0.484 | 0.737 | 0.921 | 0.12 | 0.43 | 0.729 | 0.933 | 0.31 | 0.854 | Non-time varying |
| Ovarian | 2.05 | 2.32 | 5.38 | 0.152 | 0.313 | 0.250 | 0.28 | 3.33 | 0.600 | 0.343 | 3.06 | 0.217 | Non-time varying |
| Ovarian (serous) | 0.02 | 3.82 | 6.40 | 0.886 | 0.148 | 0.171 | 3.80 | 6.38 | 0.051 | 0.094 | 2.59 | 0.274 | Non-time varying |
| Ovarian (non-serous) | 2.40 | 2.94 | 6.65 | 0.121 | 0.230 | 0.155 | 0.53 | 4.25 | 0.465 | 0.236 | 3.72 | 0.156 | Non-time varying |
| Uterine | 5.60 | 6.02 | 10.56 | **0.018** | **0.049** | **0.032** | 0.42 | 4.96 | 0.518 | 0.175 | 4.54 | 0.103 | 0 spline |
| Thyroid | 0.74 | 0.84 | 1.62 | 0.389 | 0.659 | 0.806 | 0.09 | 0.87 | 0.761 | 0.832 | 0.78 | 0.677 | Non-time varying |
| Colorectal | 1.64 | 4.09 | 5.63 | 0.200 | 0.129 | 0.229 | 2.45 | 3.99 | 0.117 | 0.263 | 1.54 | 0.464 | Non-time varying |
| Melanoma (invasive) | 0.12 | 0.12 | 5.71 | 0.725 | 0.940 | 0.222 | 0.00 | 5.59 | 0.985 | 0.134 | 5.59 | 0.061 | Non-time varying |
| Melanoma (in-situ) | 0.74 | 4.53 | 5.38 | 0.390 | 0.104 | 0.251 | 3.79 | 4.64 | 0.052 | 0.200 | 0.85 | 0.653 | Non-time varying |
| Pancreatic | 0.21 | 0.21 | 1.96 | 0.649 | 0.901 | 0.742 | 0.00 | 1.76 | 0.971 | 0.625 | 1.75 | 0.416 | Non-time varying |
| Lung | 0.41 | 0.43 | 1.41 | 0.523 | 0.808 | 0.843 | 0.02 | 1.00 | 0.890 | 0.802 | 0.98 | 0.613 | Non-time varying |
| Haematological | 0.66 | 3.24 | 7.66 | 0.418 | 0.198 | 0.105 | 2.59 | 7.00 | 0.108 | 0.072 | 4.42 | 0.110 | Non-time varying |

ART=Assisted Reproductive Technology; IUI/OS=Intrauterine Insemination/Ovarian Stimulation. NA indicates model did not fit

**Supplementary Table 8:**
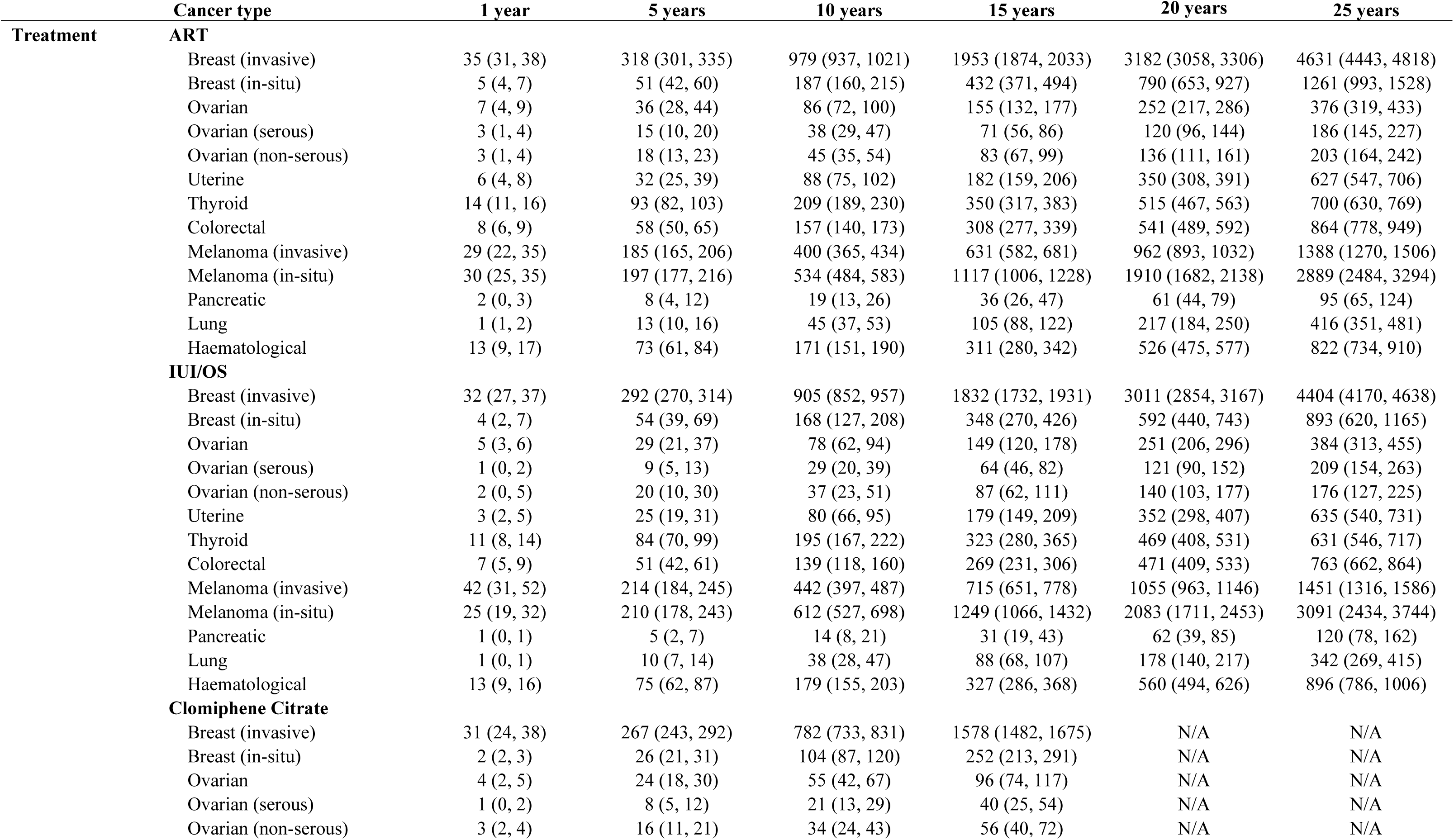
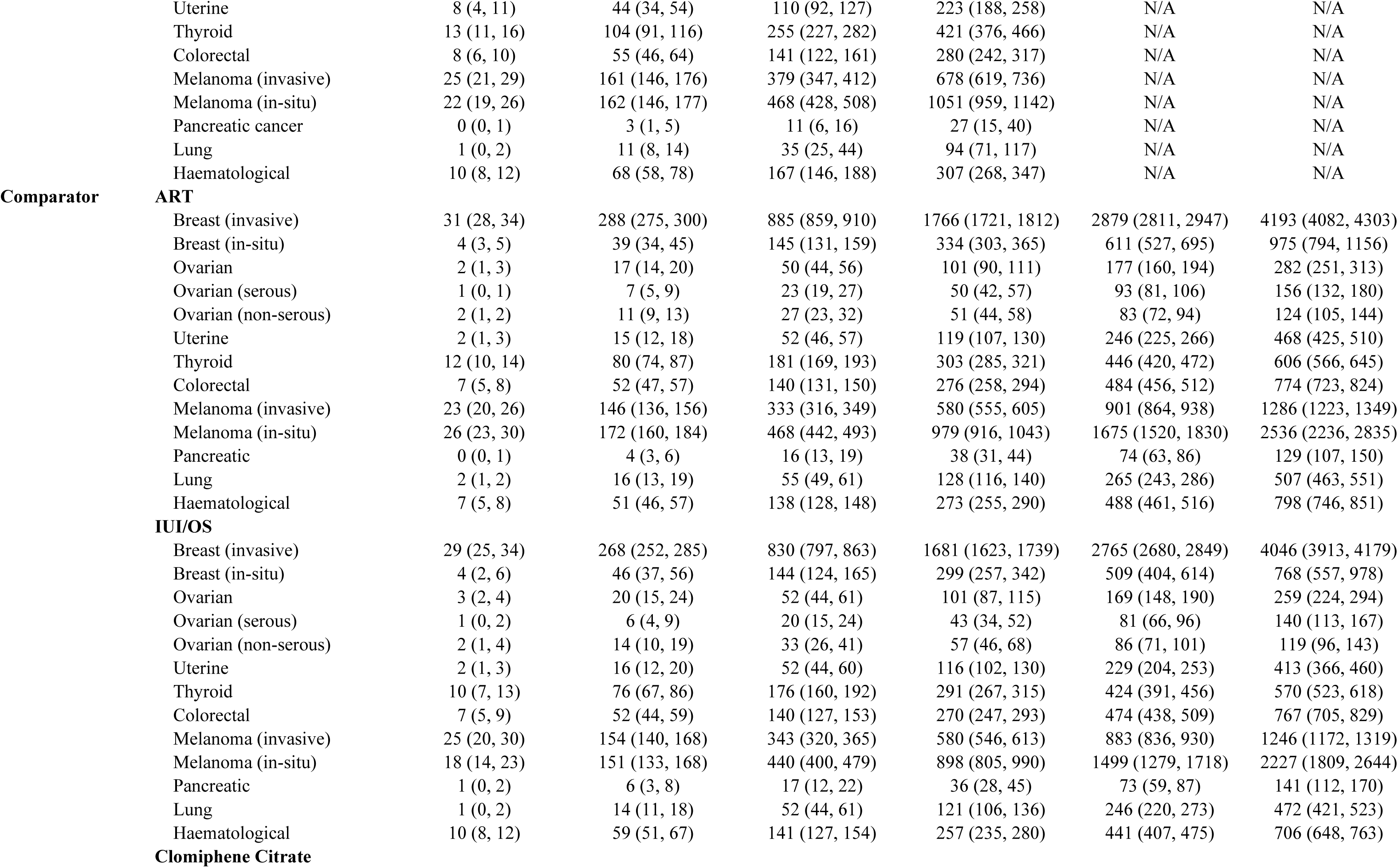
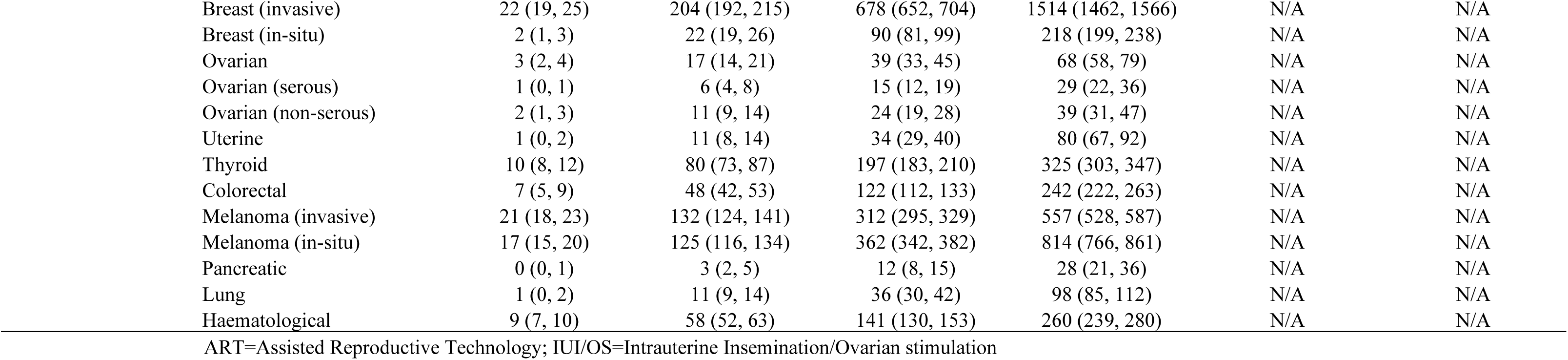
Predicted number of cancers for treatment and comparator cohorts per 100,000 women.

**Supplementary Table 9:**
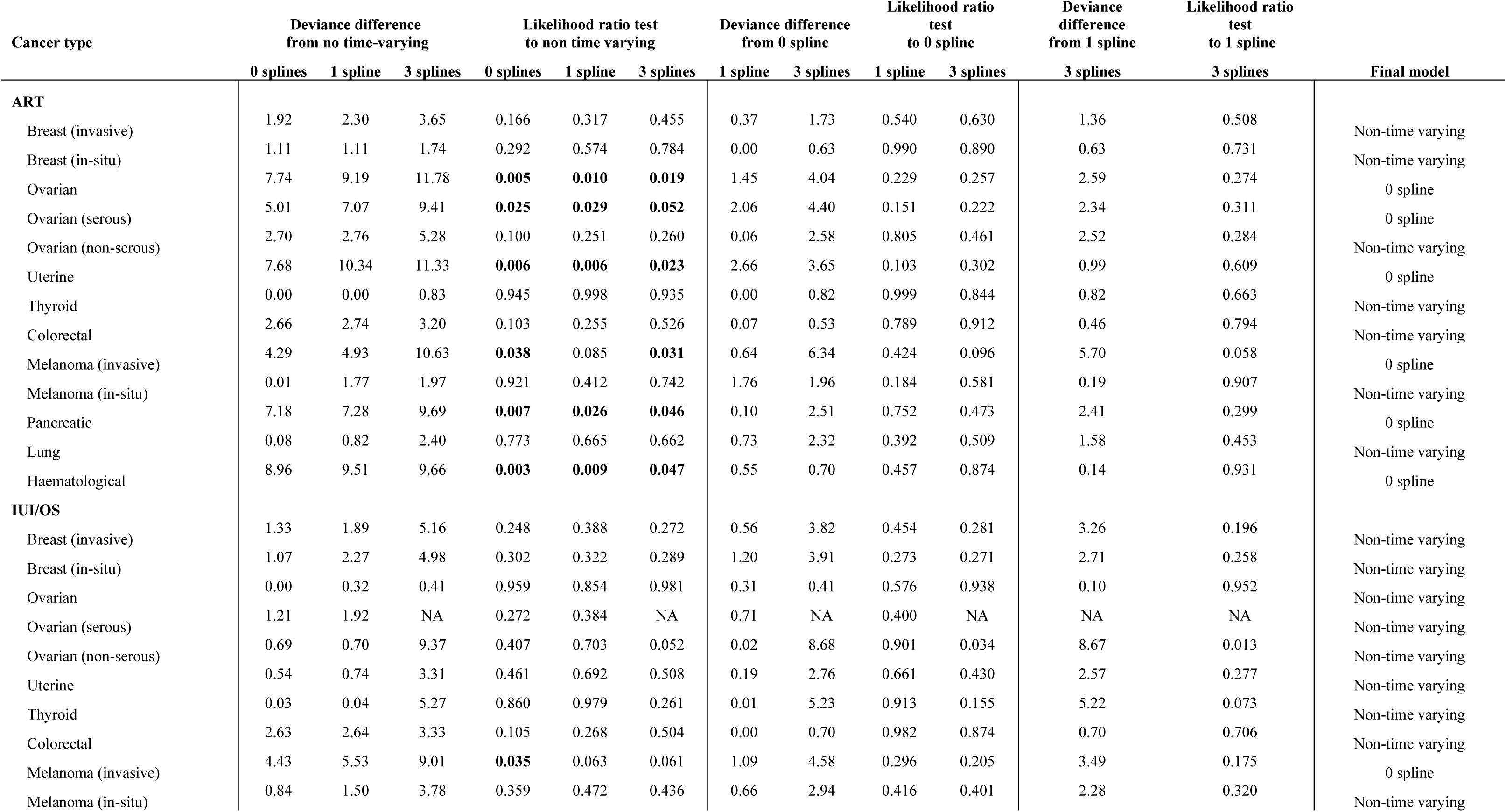
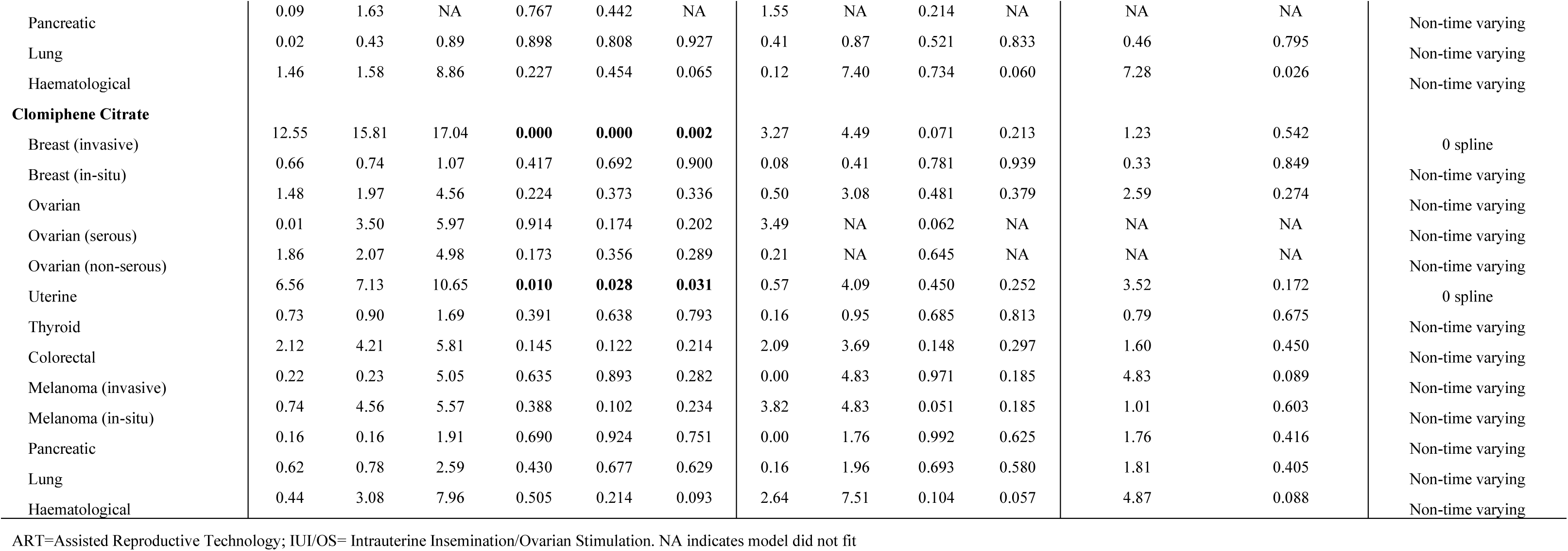
Deviance and likelihood ratio tests for selection of models (No prior cancer sensitivity analysis).

| Cancer type | Deviance difference<br>from no time-varying |  |  | Likelihood ratio test<br>to non time varying |  |  | Deviance difference<br>from 0 spline |  | Likelihood ratio<br>test<br>to 0 spline |  | Deviance<br>difference<br>from 1 spline | Likelihood ratio<br>test<br>to 1 spline | Final model |
| --- | --- | --- | --- | --- | --- | --- | --- | --- | --- | --- | --- | --- | --- |
|  | 0 splines | 1 spline | 3 splines | 0 splines | 1 spline | 3 splines | 1 spline | 3 splines | 1 spline | 3 splines | 3 splines | 3 splines |  |
| ART |  |  |  |  |  |  |  |  |  |  |  |  |  |
| Breast (invasive) | 1.92 | 2.30 | 3.65 | 0.166 | 0.317 | 0.455 | 0.37 | 1.73 | 0.540 | 0.630 | 1.36 | 0.508 | Non-time varying |
| Breast (in-situ) | 1.11 | 1.11 | 1.74 | 0.292 | 0.574 | 0.784 | 0.00 | 0.63 | 0.990 | 0.890 | 0.63 | 0.731 | Non-time varying |
| Ovarian | 7.74 | 9.19 | 11.78 | 0.005 | 0.010 | 0.019 | 1.45 | 4.04 | 0.229 | 0.257 | 2.59 | 0.274 | 0 spline |
| Ovarian (serous) | 5.01 | 7.07 | 9.41 | 0.025 | 0.029 | 0.052 | 2.06 | 4.40 | 0.151 | 0.222 | 2.34 | 0.311 | 0 spline |
| Ovarian (non-serous) | 2.70 | 2.76 | 5.28 | 0.100 | 0.251 | 0.260 | 0.06 | 2.58 | 0.805 | 0.461 | 2.52 | 0.284 | Non-time varying |
| Uterine | 7.68 | 10.34 | 11.33 | 0.006 | 0.006 | 0.023 | 2.66 | 3.65 | 0.103 | 0.302 | 0.99 | 0.609 | 0 spline |
| Thyroid | 0.00 | 0.00 | 0.83 | 0.945 | 0.998 | 0.935 | 0.00 | 0.82 | 0.999 | 0.844 | 0.82 | 0.663 | Non-time varying |
| Colorectal | 2.66 | 2.74 | 3.20 | 0.103 | 0.255 | 0.526 | 0.07 | 0.53 | 0.789 | 0.912 | 0.46 | 0.794 | Non-time varying |
| Melanoma (invasive) | 4.29 | 4.93 | 10.63 | 0.038 | 0.085 | 0.031 | 0.64 | 6.34 | 0.424 | 0.096 | 5.70 | 0.058 | 0 spline |
| Melanoma (in-situ) | 0.01 | 1.77 | 1.97 | 0.921 | 0.412 | 0.742 | 1.76 | 1.96 | 0.184 | 0.581 | 0.19 | 0.907 | Non-time varying |
| Pancreatic | 7.18 | 7.28 | 9.69 | 0.007 | 0.026 | 0.046 | 0.10 | 2.51 | 0.752 | 0.473 | 2.41 | 0.299 | 0 spline |
| Lung | 0.08 | 0.82 | 2.40 | 0.773 | 0.665 | 0.662 | 0.73 | 2.32 | 0.392 | 0.509 | 1.58 | 0.453 | Non-time varying |
| Haematological | 8.96 | 9.51 | 9.66 | 0.003 | 0.009 | 0.047 | 0.55 | 0.70 | 0.457 | 0.874 | 0.14 | 0.931 | 0 spline |
| IUI/OS |  |  |  |  |  |  |  |  |  |  |  |  |  |
| Breast (invasive) | 1.33 | 1.89 | 5.16 | 0.248 | 0.388 | 0.272 | 0.56 | 3.82 | 0.454 | 0.281 | 3.26 | 0.196 | Non-time varying |
| Breast (in-situ) | 1.07 | 2.27 | 4.98 | 0.302 | 0.322 | 0.289 | 1.20 | 3.91 | 0.273 | 0.271 | 2.71 | 0.258 | Non-time varying |
| Ovarian | 0.00 | 0.32 | 0.41 | 0.959 | 0.854 | 0.981 | 0.31 | 0.41 | 0.576 | 0.938 | 0.10 | 0.952 | Non-time varying |
| Ovarian (serous) | 1.21 | 1.92 | NA | 0.272 | 0.384 | NA | 0.71 | NA | 0.400 | NA | NA | NA | Non-time varying |
| Ovarian (non-serous) | 0.69 | 0.70 | 9.37 | 0.407 | 0.703 | 0.052 | 0.02 | 8.68 | 0.901 | 0.034 | 8.67 | 0.013 | Non-time varying |
| Uterine | 0.54 | 0.74 | 3.31 | 0.461 | 0.692 | 0.508 | 0.19 | 2.76 | 0.661 | 0.430 | 2.57 | 0.277 | Non-time varying |
| Thyroid | 0.03 | 0.04 | 5.27 | 0.860 | 0.979 | 0.261 | 0.01 | 5.23 | 0.913 | 0.155 | 5.22 | 0.073 | Non-time varying |
| Colorectal | 2.63 | 2.64 | 3.33 | 0.105 | 0.268 | 0.504 | 0.00 | 0.70 | 0.982 | 0.874 | 0.70 | 0.706 | Non-time varying |
| Melanoma (invasive) | 4.43 | 5.53 | 9.01 | 0.035 | 0.063 | 0.061 | 1.09 | 4.58 | 0.296 | 0.205 | 3.49 | 0.175 | 0 spline |
| Melanoma (in-situ) | 0.84 | 1.50 | 3.78 | 0.359 | 0.472 | 0.436 | 0.66 | 2.94 | 0.416 | 0.401 | 2.28 | 0.320 | Non-time varying |
| Pancreatic | 0.09 | 1.63 | NA | 0.767 | 0.442 | NA | 1.55 | NA | 0.214 | NA | NA | NA | Non-time varying |
| Lung | 0.02 | 0.43 | 0.89 | 0.898 | 0.808 | 0.927 | 0.41 | 0.87 | 0.521 | 0.833 | 0.46 | 0.795 | Non-time varying |
| Haematological | 1.46 | 1.58 | 8.86 | 0.227 | 0.454 | 0.065 | 0.12 | 7.40 | 0.734 | 0.060 | 7.28 | 0.026 | Non-time varying |
| **Clomiphene Citrate** |  |  |  |  |  |  |  |  |  |  |  |  |  |
| Breast (invasive) | 12.55 | 15.81 | 17.04 | **0.000** | **0.000** | **0.002** | 3.27 | 4.49 | 0.071 | 0.213 | 1.23 | 0.542 | 0 spline |
| Breast (in-situ) | 0.66 | 0.74 | 1.07 | 0.417 | 0.692 | 0.900 | 0.08 | 0.41 | 0.781 | 0.939 | 0.33 | 0.849 | Non-time varying |
| Ovarian | 1.48 | 1.97 | 4.56 | 0.224 | 0.373 | 0.336 | 0.50 | 3.08 | 0.481 | 0.379 | 2.59 | 0.274 | Non-time varying |
| Ovarian (serous) | 0.01 | 3.50 | 5.97 | 0.914 | 0.174 | 0.202 | 3.49 | NA | 0.062 | NA | NA | NA | Non-time varying |
| Ovarian (non-serous) | 1.86 | 2.07 | 4.98 | 0.173 | 0.356 | 0.289 | 0.21 | NA | 0.645 | NA | NA | NA | Non-time varying |
| Uterine | 6.56 | 7.13 | 10.65 | **0.010** | **0.028** | **0.031** | 0.57 | 4.09 | 0.450 | 0.252 | 3.52 | 0.172 | 0 spline |
| Thyroid | 0.73 | 0.90 | 1.69 | 0.391 | 0.638 | 0.793 | 0.16 | 0.95 | 0.685 | 0.813 | 0.79 | 0.675 | Non-time varying |
| Colorectal | 2.12 | 4.21 | 5.81 | 0.145 | 0.122 | 0.214 | 2.09 | 3.69 | 0.148 | 0.297 | 1.60 | 0.450 | Non-time varying |
| Melanoma (invasive) | 0.22 | 0.23 | 5.05 | 0.635 | 0.893 | 0.282 | 0.00 | 4.83 | 0.971 | 0.185 | 4.83 | 0.089 | Non-time varying |
| Melanoma (in-situ) | 0.74 | 4.56 | 5.57 | 0.388 | 0.102 | 0.234 | 3.82 | 4.83 | 0.051 | 0.185 | 1.01 | 0.603 | Non-time varying |
| Pancreatic | 0.16 | 0.16 | 1.91 | 0.690 | 0.924 | 0.751 | 0.00 | 1.76 | 0.992 | 0.625 | 1.76 | 0.416 | Non-time varying |
| Lung | 0.62 | 0.78 | 2.59 | 0.430 | 0.677 | 0.629 | 0.16 | 1.96 | 0.693 | 0.580 | 1.81 | 0.405 | Non-time varying |
| Haematological | 0.44 | 3.08 | 7.96 | 0.505 | 0.214 | 0.093 | 2.64 | 7.51 | 0.104 | 0.057 | 4.87 | 0.088 | Non-time varying |

ART=Assisted Reproductive Technology; IUI/OS= Intrauterine Insemination/Ovarian Stimulation. NA indicates model did not fit

**Supplementary Table 10:**
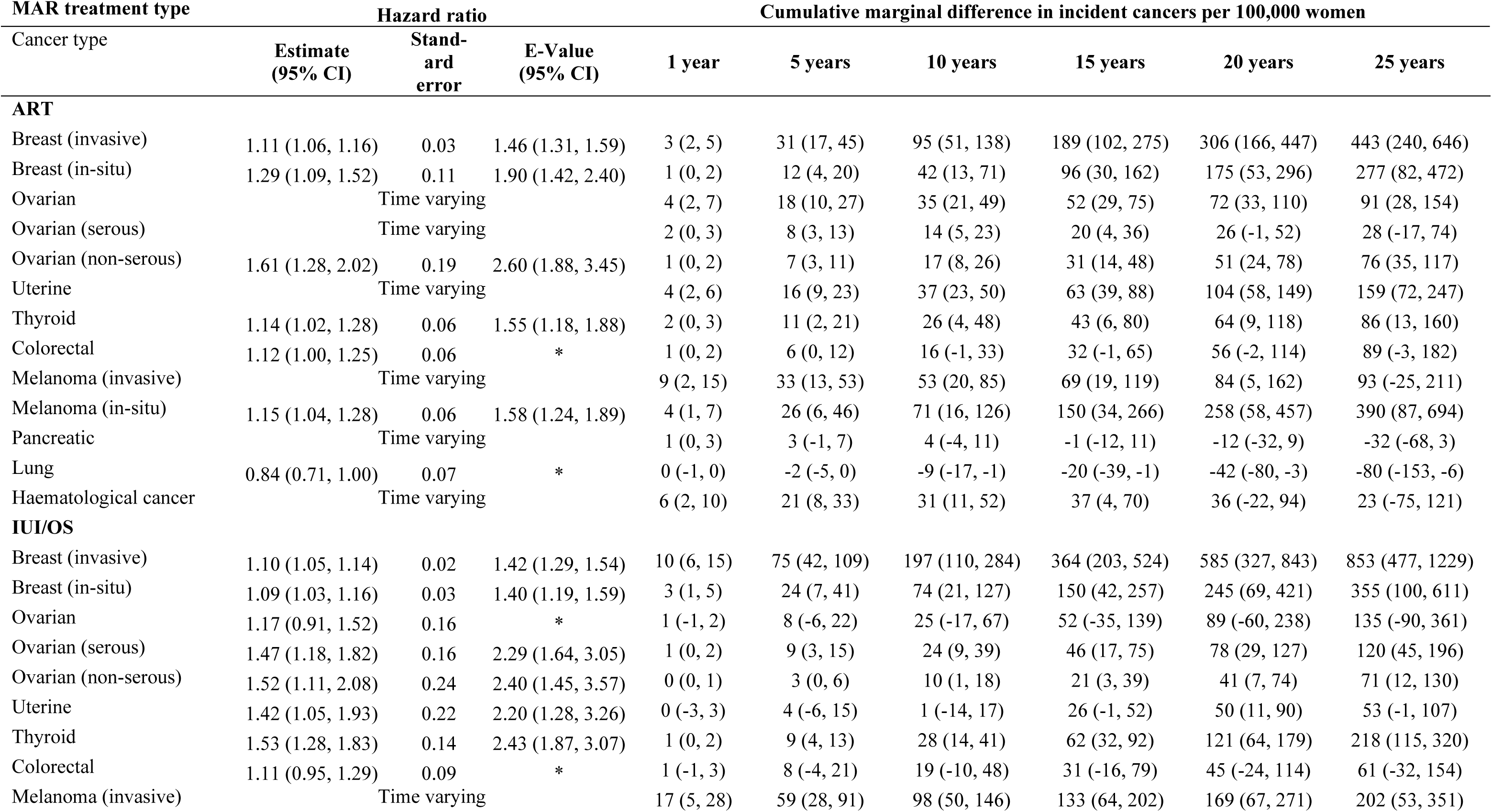
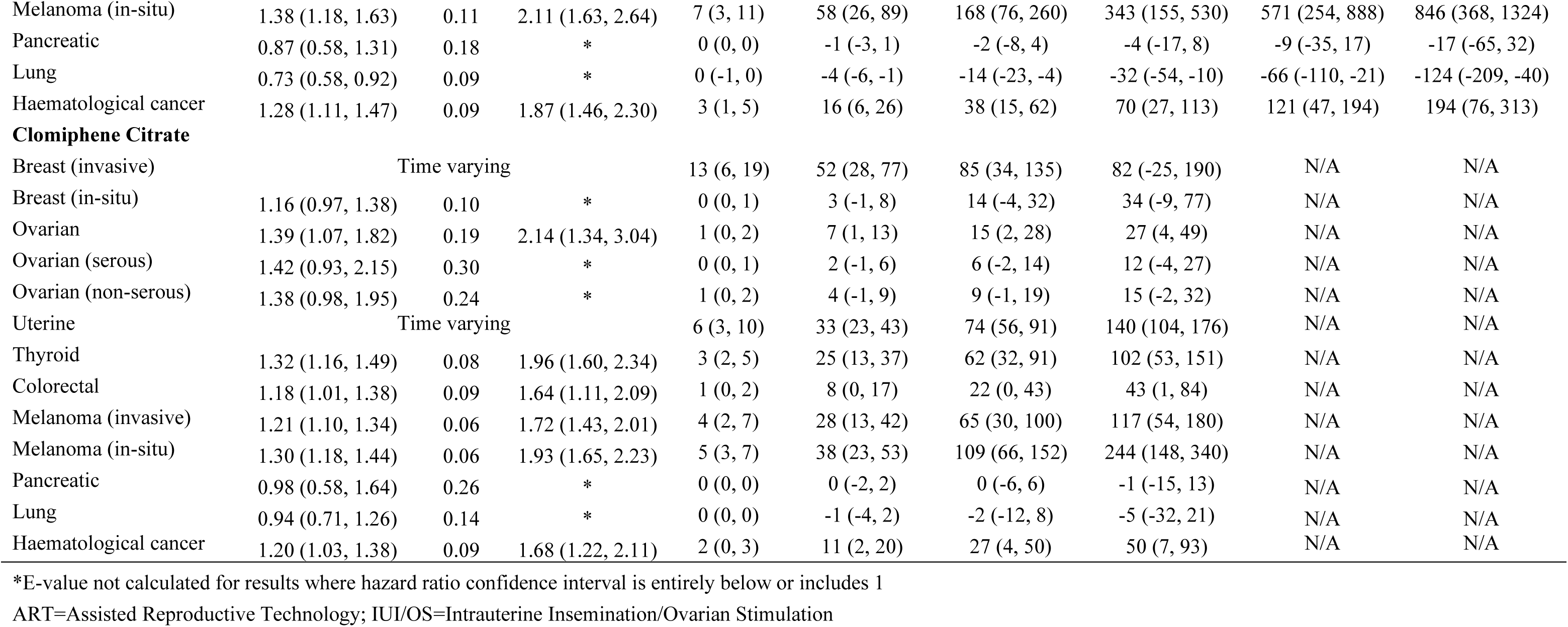
Hazard ratios, E-values, and cumulative marginal difference in incident cancers (per 100,000 women) for each emulated t trial (No prior cancer sensitivity analysis).

**Supplementary Table 11:**
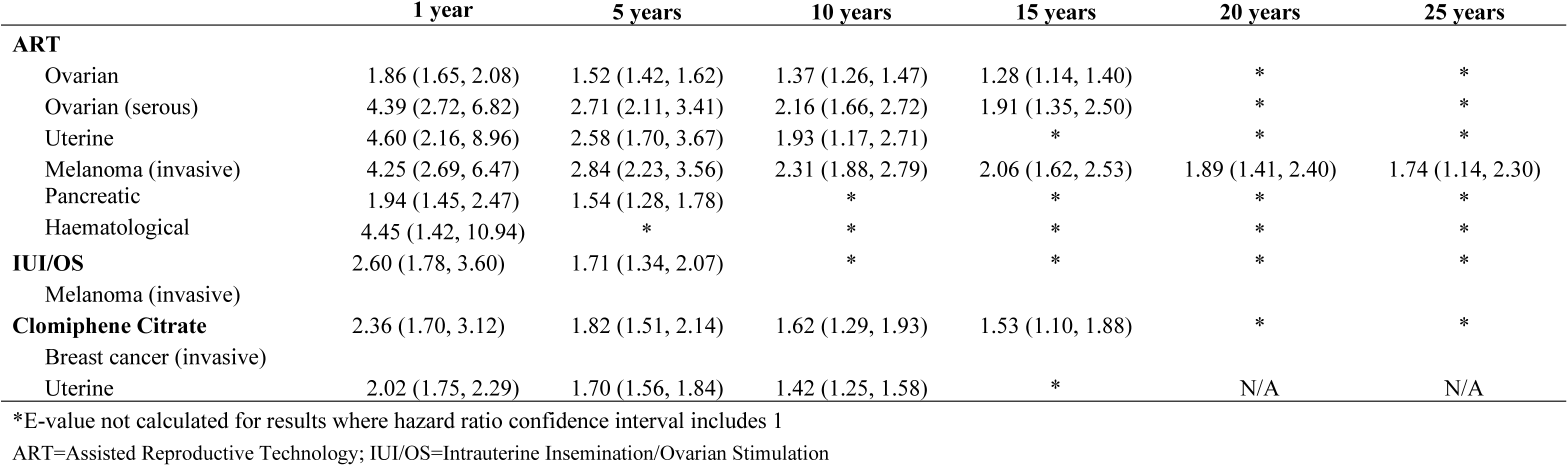
E-values over time for time-varying hazard ratio models (No prior cancer sensitivity analysis)

## 15. SUPPLEMENTARY FIGURES

**Supplementary Figure 1:**
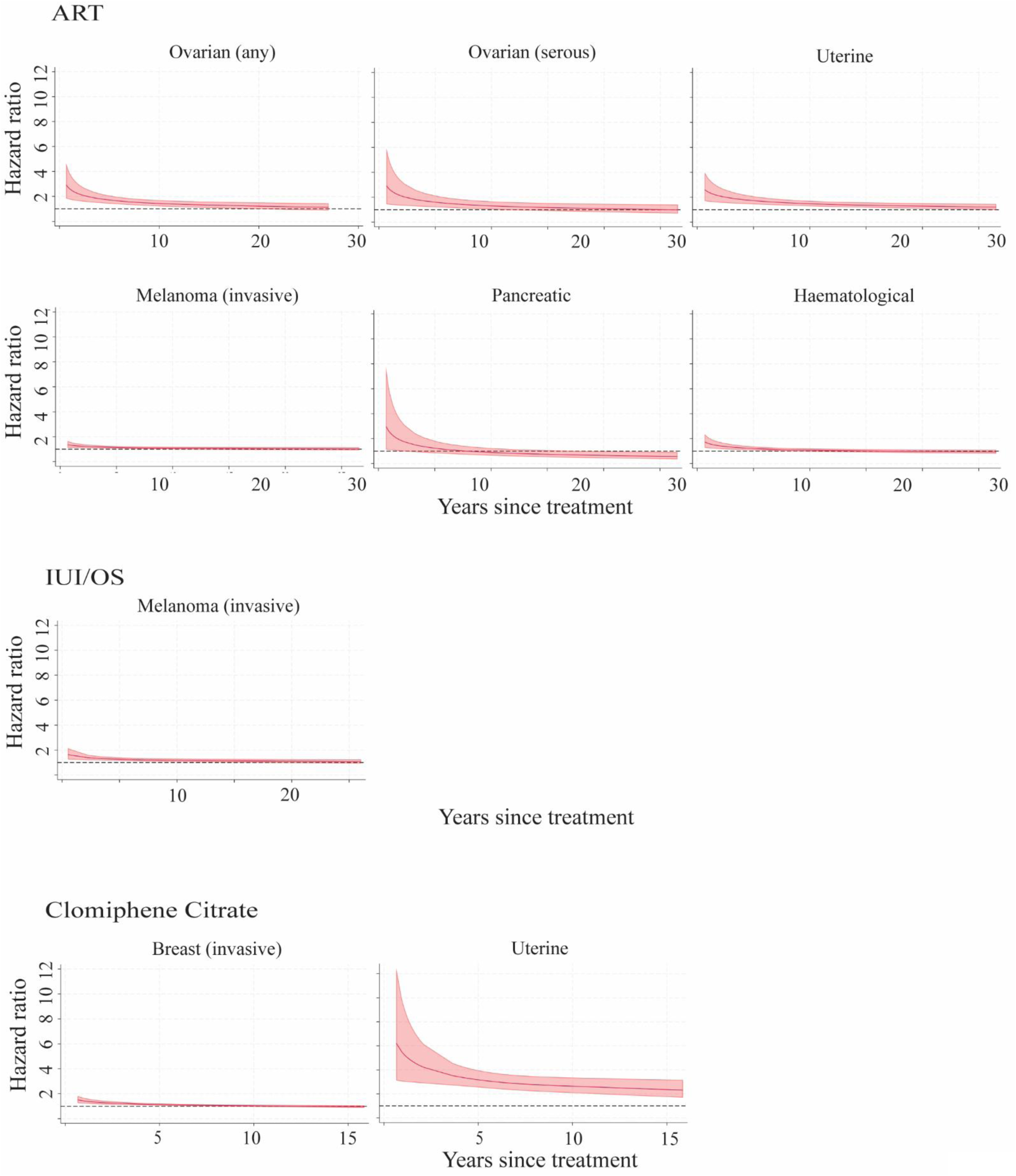
Hazard ratios over time for cancers with time-varying associations with treatments (No prior cancer sensitivity analysis). Figures start from 6 months to improve figure ability.

